# Safety, Pharmacokinetics, and Cardiodynamics of CMS121, a Novel Small Molecule Fisetin Derivative with Neuroprotective Properties, in Phase 1 Healthy Adult Volunteers

**DOI:** 10.1101/2025.02.28.25323123

**Authors:** Pamela Maher, Ronald Christopher, Rebecca Evans, William Raschke

## Abstract

The safety, tolerability, pharmacokinetics, age-related effects of single (SD) and repeat (RD) doses of CMS121, a novel small molecule fisetin derivative, were evaluated in healthy adult volunteers. The effects of food were also evaluated in healthy young adult subjects. SD of up to 1800 mg or RD up to 900 mg/day for 7 days was generally well tolerated, with the majority of TEAEs mild in severity. Generally, the pharmacokinetics of CMS121 and its metabolites were well characterized and increased in a dose-proportional or slightly greater than dose-proportional manner across the range of doses assessed. CMS121-C2 metabolite appears to contribute the most to the presence of the molar-equivalent CMS121 in plasma than the parent compound or the other metabolites (i.e. CMS121-C1 and CMS121-C3). Urinary excretion of CMS121 metabolites was minimal, implying urinary excretion may not be a major clearance route by which CMS121 is eliminated after oral dosing. There is a significant effect of age on the pharmacokinetics of CMS121 and its metabolites, with higher systemic exposures to CMS121 and its metabolites and longer terminal elimination half-lives in elderly subjects. Systemic exposures to CMS121 were higher in the fed state by approximately 50%.

---

Alzheimer’s disease (AD) is the most frequent age-associated disease with a lack of effective treatments to prevent, delay, slow or stop the cognitive and other functional changes that are the clinical hallmarks of the disease [1, 2]. The greatest known risk factor for AD is increasing age [3, 4]. About 1 in 9 people age 65 and older has AD, and the prevalence is 1 in 3 people over the age of 85 [4]. These data suggest that degenerative processes associated with old age offer excellent targets for treating AD. A novel approach for AD treatment that uses cell-based phenotypic screening assays was developed to identify compounds that protect neurons from a variety of age-associated toxicities including ferroptosis, in vitro ischemia, inflammation, trophic factor loss and proteotoxicity [5-7]. Using these assays, we initially identified the flavonoid fisetin (3,7,3’,4’ tetrahydroxyflavone) as an orally active, novel neuroprotective and cognition-enhancing molecule (Maher, 2021). Fisetin has both direct antioxidant activity and maintains the levels of GSH under conditions of stress by inducing the transcription factors, Nrf2 and ATF4 (Ehren & Maher, 2013). Moreover, fisetin was shown to facilitate long term potentiation both in hippocampal slices (Maher et al., 2006) and in mouse brains (He et al., 2018) and oral administration of fisetin promoted learning and memory in mice using the object recognition test (Maher et al., 2006). Animal studies showed that fisetin prevented learning and memory deficits in APPswe/PS1dE9 (huAPP/PS1) double transgenic AD mice (Currais et al., 2014) and rapidly aging SAMP8 mice (Currais et al., 2018). Fisetin was also effective in two different models of stroke (Gelderblom et al., 2012; Maher et al., 2007) and three different models of Huntington’s disease (Maher et al., 2011).

CMS121 is a derivative of fisetin (Chiruta et al., 2012) (Figure 1). Using structure-activity relationship-driven iterative chemistry, we synthesized more than 160 derivatives of fisetin based on several different chemical scaffolds. We then used a multi-tiered approach to screening that allowed us to identify fisetin derivatives with significantly enhanced neuroprotective activity in two in vitro neuroprotection assays while at the same time maintaining other key actions including anti-inflammatory activity. While all of the fisetin derivatives had improved medicinal chemical properties, more consistent with those of known CNS drugs, ∼20 of them had greatly enhanced neuroprotective activity. Absorption, distribution, metabolism, and excretion and PK studies on the six most promising derivatives showed that several had peak brain levels following a single oral dose of 20 mg/kg that greatly exceeded their average median effective concentration in the in vitro neuroprotection assays as well as good oral bioavailability. Based on these and other results, we selected two fisetin derivatives (CMS121 and CMS140) to test in transgenic AD mice for their ability to prevent cognitive dysfunction. The results from these behavioral assays showed that while both CMS140 and CMS121 can rescue the cognitive dysfunction associated with AD when administered at a late stage in the disease process when pathology is already present, CMS121 was significantly more effective than CMS140 (Gamze Ates et al, 2020). Thus, further work focused on the development of CMS121 for the treatment of AD.

**Figure 1.**
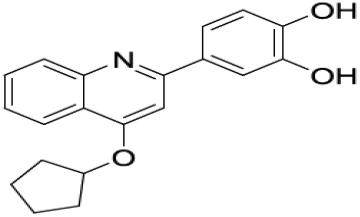
Chemical structure of CMS121

Further studies identified the target of CMS121 as fatty acid synthase (FASN) which is partially inhibited by the compound (Gamze Ates et al, 2020). Inhibition of FASN by CMS121 results in reductions in both lipid peroxidation and inflammation, both of which are thought to play key roles in the cognitive dysfunction that it is the hallmark of AD (Bradley-Whitman MA et al., 2015; Reed T, 2011; Rukhsana S. et al. 2013; Jefferson WK et al. 2018; Heneka MT, et.al, 2024). Thus, CMS121 targets multiple changes that are implicated in the development and progression of AD and therefore holds great promise for the treatment of the disease.

This manuscript summarizes data from Phase 1 human studies designed to assess the safety, tolerability, and PK of single and multiple ascending doses of CMS121 in healthy young adult subjects (Parts 1 and 2) and the safety, tolerability, and PK profile of multiple doses of CMS121 in healthy elderly subjects (NCT05318040). Additionally, the effect of food on the rate and extent of absorption of CMS121 was also investigated when administered to healthy young adult subjects after a period of fasting and shortly after a high-fat/high-calorie meal.

## Methods

### Study Population

Healthy, adult, male or female volunteers, 19-60 years of age, inclusive, and for studies in elderly, healthy male or female, 65-85 years of age, inclusive were recruited in the studies. Eligibility was based on no abnormalities in past medical history, physical examination, vital signs, clinical laboratory parameters, and electrocardiography (ECG). All relevent ethical guidelines were followed including IRB approval prior to study initiation. All subjects gave written informed consent prior to their participation in the study. Concomitant medications were restricted in accordance with protocol inclusion and exclusion criteria. All doses of CMS121/placebo were administered orally with approximately 240 mL of water. For doses consisting of ≥ 4 capsules to swallow, additional water, up to a maximum of 50 mL, was administered as required by the subject, however, all dosing was completed within 5 minutes.

### Study Design

This was a 4-part study. Parts 1, 2, and 3 were randomized, double-blind, placebo-controlled investigations of CMS121 administered orally (po) with food. First, single ascending doses were administered in healthy young adult subjects (SAD: Part 1, 50 to 1800 mg po once). Then, multiple ascending doses were administered in healthy young adult subjects (MAD: Part 2, 150 to 900 mg po once daily [QD] for 7 days), and multiple doses were administered in healthy elderly subjects (Part 3, 600 mg po QD for 7 days). Part 4 was an open-label, 2-way crossover study to assess the effect of food on single oral doses of CMS121 (600 mg po QD once after a period of fasting and 600 mg po QD once after a high-fat/high-calorie meal) in healthy young adult subjects.

Up to 100 healthy, adult male and female subjects were planned to be enrolled. Subjects participated in only 1 study part and in only 1 cohort.

### Drug Formulation and Administration

CMS121 was supplied as a CMS121 maleate salt form powder in capsules. Placebo (microcrystalline cellulose) was provided as visually matching placebo capsules.

### Clinical Assessments

In each of the four studies, each subject was screened for eligibility within 28 days prior to dosing, examined on Day-1 for baseline physical and laboratory assessments, and monitored for safety and tolerability endpoints after dosing at protocol defined internals until the end of follow up. Safety parameters, including physical examinations, vital signs, clinical laboratory parameters (including blood biochemistry, hematology, and urinalysis), 12-lead safety ECGs, and C-SSRS, were assessed throughout the study.

Plasma and urine samples were collected predose and through 72 hours following CMS121 administration for the PK assessments of CMS121 and its metabolites. ECGs were collected predose and up to 24 hours following Day 1 dosing.

Interim plasma PK analyses was performed to evaluate appropriate sampling time points as the single dose phase progressed. Interim plasma PK analyses were performed prior to conducting Parts 2 (MAD), 3 (Elderly) and 4 (Food effect).

#### Part 1 (SAD)

Up to 6 cohorts of 8 subjects each (6 active and 2 placebo) were planned for evaluation. The first cohort, Cohort S1, included a sentinel group (1 active and 1 placebo) which was dosed at least 24 hours before the remaining 6 subjects (5 active and 1 placebo). Dosing of the remaining 6 subjects was conducted following a safety evaluation of the sentinel group by the safety review committee (SRC). Dose escalation to the next dose level (i.e., next cohort) did not take place until the safety review committee (SRC) had determined that adequate safety and tolerability were demonstrated in previous cohort(s) to permit proceeding to the next cohort.

In each cohort, subjects received a single oral dose of CMS121 (50 to 1800 mg) or placebo under fed conditions.

#### Part 2 (MAD)

Part 2 started following the completion of Part 1. Up to 4 cohorts of 8 subjects each (6 active and 2 placebo) were planned for evaluation. A decision not to include sentinel dosing was made based on safety, tolerability, and plasma PK data from Part 1. Dose escalation to the next dose level (i.e., next cohort) did not take place until the SRC had determined that adequate safety and tolerability were demonstrated in previous cohort(s) to permit proceeding to the next cohort.

In each cohort, subjects received multiple oral doses of CMS121 (150 to 900 mg) or placebo QD for 7 days under fed conditions. Plasma samples were collected predose on Day 1, through 24 hours following CMS121 administration on Day 1, and through 72 hours following CMS121 administration on Day 7 for the PK assessments of CMS121 and its metabolites. Plasma predose samples were also collected in the morning of Days 5, 6, and 7. Following review of the urine PK data in Part 1, urine samples were collected predose on Day 1 and through 72 hours following CMS121 administration on Day 7 for the PK assessments. of CMS121 and its metabolites. Interim plasma PK analyses were performed prior to conducting Part 3.

ECGs were collected predose on Days 1 and 7, and up to 24 hours following Day 7 dosing.

#### Part 3 (Elderly)

Initiation of Part 3 started following the completion of Parts 1 and 2. Initiation of Part 3 did not take place until the SRC had determined that adequate safety, tolerability, and plasma PK were demonstrated in Part 2 for at least one level higher than the planned dose level in Part 3 and all previously conducted cohorts in Parts 1 and 2.

One (1) cohort of 8 subjects (6 active and 2 placebo) was planned for evaluation. Subjects received multiple oral doses of CMS121 (600 mg) or placebo QD for 7 days under fed conditions.

Plasma samples were collected predose on Day 1, through 24 hours following CMS121 administration on Day 1, and through 72 hours following CMS121 administration on Day 7 for the PK assessments of CMS121 and its metabolites. Plasma predose samples were also collected in the morning of Days 5, 6, and 7. Urine samples were collected predose on Day 1 and through 72 hours following CMS121 administration on Day 7 for the PK assessments of CMS121 and its metabolites.

#### Part 4 (Food Effect)

Initiation of Part 4 started following the completion of Parts 1-3. Twelve subjects were planned for this evaluation. On Day 1 of each of 2 treatment periods, a single oral dose of CMS121 (600 mg) was administered following either a standard high-fat/high-calorie meal (Treatment A) or an overnight fast (Treatment B) as per each subject’s assigned randomization sequence (AB or BA). Plasma samples were collected predose and through 72 hours following CMS121 administration for the PK assessments of CMS121 and its metabolites. There was a washout of at least 7 days between doses.

### Pharmacokinetic Assessments

Blood samples were collected into (K_2_EDTA) tubes. Immediately after collection, the blood was gently inverted to mix with the anticoagulant and placed on ice or in a refrigerator. The tube was gently mixed, plasma fractions separated and then stored at -20°C until analysis.

Sample analyses for CMS121 and its glucuronide conjugate metabolites (e.g., CMS121-C1, CMS121-C2, and CMS121-C3) in plasma and urine were performed at Celerion, Inc. Plasma concentrations of CMS121 and its metabolites, CMS121-C1, CMS121-C2, and CMS121-C3, as determined per the bioanalytical method and the collection times, were used for the calculation of the plasma PK parameters.

The appropriate noncompartmental PK parameters were calculated from the plasma CMS121, CMS121-C1, CMS121-C2, and CMS121-C3 concentration-time data using Phoenix^®^ WinNonlin^®^ Version 8.3.4. Actual sample times were used in the calculations of the PK parameters. The calculation of the actual time was in respect to the dose administration time of CMS121 on Day 1 (all study parts) and Day 7 (MAD and Elderly only).

Determination of CMS121 in human plasma was performed over the range 0.05–25 ng/mL using validated HPLC-MS/MS. Similarly, quantification of metabolites CMS121-1 and CMS121-2 in plasma was performed over the range of 0.25–125 ng/mL and for CMS121-3 over the range of 10–2000 ng/mL using validated LC-MS/MS methodology. CMS121 and metabolites were processed from plasma fractions by protein precipitation using acetonitrile containing deuterated internal standards (d_7_-CMS121 (IS), d_7_-CMS121-C2 and d_7_-CMS121-C3 (IS)). Stability in human plasma (K_2_/EDTA) at room temperature was 24 hr and at -20°C up to 258 days.

### ECG Assessments

For study conduct, ECGs were classified as either 12-lead Safety ECGs, which were collected for real-time safety review, or 12-lead Cardiodynamic ECGs, which were extracted from the Holter monitor recordings. Safety 12-lead ECGs were not used for inferential analysis.

Holter monitors were used to collect continuous 12-lead ECG data for the purpose of extracting ECG parameters. Triplicate 10-second, 12-lead ECG recordings were extracted from the Holter monitor data within a 5-minute time window ending approximately 1-2 minutes prior to the corresponding PK blood sample collection around the following time points relative to dosing (Table 1; Cardiodynamics: 12-lead ECG Holter Monitor Extraction Time Points; see Supplemental).

**Table 1:** Cardiodynamics: 12-lead ECG Holter Monitor Extraction Time Points

| Study Part | ECG Parameter | Day | Time Points |
| --- | --- | --- | --- |
| SAD | HR, RR, PR, QRS, QT, QTcF | 1 | Predose (mean of -60, -45, and -30 minutes)<br>Hours 0.25, 0.5, 1, 1.5, 2, 3, 4, 6, 8, and 12 |
|  |  | 2 | Hour 24 |
| MAD | HR, RR, PR, QRS, QT, QTcF | 1 | Predose (mean of -60, -45, and -30 minutes) |
|  |  | 7 | Predose (-30 minutes)<br>Hours 0.25, 0.5, 1, 1.5, 2, 3, 4, 6, 8, and 12 |
|  |  | 8 | Hour 24 |

Attempts were made to obtain 3 interpretable ECG recordings (e.g., without artifacts) at each time point (ideally within 5 minutes of each other). The average of the 3 readings was used to determine ECG parameters (e.g., HR, RR interval, PR interval, QRS duration, QT interval, and QTcF) at each time point and was rounded to the nearest integer. These values were used in the summarizations of ECG parameters.

### Pharmacokinetic Data Analysis

All PK concentrations and PK parameter descriptive statistics were generated using SAS^®^ Version 9.4. The plasma concentrations of CMS121 and its metabolites were quantified and summarized by treatment, study day, and time point for all subjects in the PK Concentration Population. Plasma concentrations of CMS121 and its metabolites are presented with the same level of precision as received from the bioanalytical laboratory. While plasma concentrations of metabolites CMS121-C1 and CMS121-C3 were quantifiable, the levels were generally much lower across dose groups as compared with CMS121 and CMS21-C2 but followed similar concentration vs time course profiles as with the parent and CMS121-C2 metabolite. As such, only tabular listings and illustrations for CMS121 and CMS121-C2 are included in this manuscript. Summary statistics, including sample size (n), arithmetic mean (mean), standard deviation (SD), coefficient of variation (CV%), standard error of the mean (SEM), minimum, median, and maximum, were calculated for all nominal concentration time points. Excluded subjects were included in the concentration listings, but were excluded from the summary statistics and noted as such in the tables. All BLQ values were presented as “BLQ” in the concentration listings and tables.

Plasma mean and individual concentration-time profiles are presented on linear and semi-log scales. Individual concentration-time profiles are based on actual sample times, and mean concentration-time profiles are based on nominal sample times. When there were significant time deviations from nominal sample time points, some concentrations were excluded from the summary statistics and any corresponding summary figures.

Plasma PK parameters, including individual treatment ratios (for Part 4), are listed and summarized by treatment and study day for all subjects in the PK Evaluable Population. PK parameters are reported to three significant figures for individual parameters, with the exception of Cmax, Clast, Cmax,ss, Ctrough, Cavg, and Cmin,ss, which are presented with same level of precision as received from the bioanalytical laboratory and Tmax, Tlag, Tmax,ss, and ratios, which are presented with two decimal places. Summary statistics (n, mean, SD, CV%, SEM, minimum, median, maximum, geometric mean [Geom Mean], and geometric CV% [Geom CV%]) are presented for all PK parameters. Excluded subjects are listed in the PK parameter tables, but are excluded from the summary statistics and noted as such in the tables.

C_trough_ values are displayed relative to the last dose given before C_trough_ (i.e., nominal time point [Day/Time]) was collected) and are presented in a separate plot from the concentration-time profiles and only on linear scale.

Urine metabolite concentrations, volumes, and PK parameters are listed and summarized by treatment, study day, and collection intervals using descriptive statistics (n, mean, SD, CV%, SEM, minimum, median, and maximum) for the PK Evaluable Population. Urine concentrations of the metabolites are presented with the same level of precision as received from the bioanalytical laboratory. Excluded subjects are listed in the urine PK tables, but are excluded from the summary statistics and noted as such in the tables. Urine concentrations that were BLQ are presented as “BLQ” in the listings and treated as zero (0) for the calculation of summary statistics. The parent CMS121 was not quantified in urine.

### Statistical Analysis of Pharmacokinetic Data

#### Dose Proportionality (Parts 1 and 2)

Dose proportionality was evaluated for CMS121 AUC0-24, AUC0-t, AUC0-inf, and Cmax on Day 1 of Part 1, AUC0-24, AUC0-t, and Cmax on Day 1 of Part 2, and AUCtau and Cmax,ss on Day 7 of Part 2 using a regression approach. Dose proportionality was assessed in 2 models: 1) SAD combined with MAD Day 1, and 2) MAD Day 7. AUC0-inf was only assessed in the SAD cohorts.

A linear relationship between the ln-transformed AUCs or Cmax and the ln-transformed dose was fitted using the following regression model:

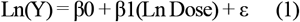

where Y represents the PK parameter. This approach is usually referred to as a power model because after exponentiation:

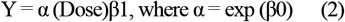

where α only depends on β0 and error. Dose proportionality required that β1 = 1 for dose-dependent parameters.

A linear relationship between the ln-transformed PK parameters and the ln-transformed dose was verified by including the cubic [ln(dose)]3 (Step 1) and quadratic [ln(dose)]2 (Step 2) terms in Model (1) using the following regression models:

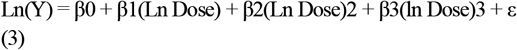

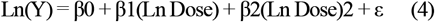

A 5% level of significance was used in sequential testing of the cubic and quadratic effects. If β2 and β3 were not statistically significantly different from 0 in Model (3) and β2 was not statistically significantly different from 0 in Model (4), a linear relationship was concluded. If any effects were statistically significant but of small magnitude (i.e., not clinically relevant), a linear relationship was concluded.

Due to the complex nature of the association between the systemic exposure and the dose, the assessment of dose proportionality was not based solely on a strict statistical rule given the small sample size for each dose level. Rather, several considerations were taken into account to assess dose proportionality including results derived from the above power model statistical analysis (i.e., the presence of a linear relationship, the slope estimates, width of the 95% CIs for the slope estimates and whether these 95% CIs contained the value of 1). In addition, qualitative assessment specific to the PK of the drug and clinical relevance was considered.

#### Steady-State Analysis (Parts 2 and 3)

A steady-state analysis was performed on the ln-transformed C_trough_ values for plasma CMS121 and metabolites for the multiple-dose cohorts. The C_trough_ values following dosing on Days 4 through 7 were used. An analysis of variance (ANOVA) model was performed separately for each treatment (Treatments M1, M2, M3, M4, and E) and included Day as a fixed effect. The best variance-covariance matrix structure was chosen to model the correlation within each subject using the Akaike’s Burnham and Anderson criterion (AICC, smaller is better).

#### Age Effect Analysis (Parts 2 and 3)

Age effect was assessed between Cohort E in Part 3 and the respective cohort (M3) in Part 2 via comparison of natural-log (ln) transformed CMS121 PK parameters AUC0-t, AUC0-inf, and C_max_ by performing an one-way ANOVA model using SAS^®^ PROC MIXED with group as a fixed effect and using the young group as the reference.

#### Food Effect Analysis (Part 4)

A comparison between Treatments A (Fed) and B (Fasted) was done via comparison of ln-transformed CMS121 PK parameters AUC0-t, AUC0-inf, and Cmax by performing an ANOVA model using SAS^®^ PROC MIXED. The ANOVA model included treatment, period, and sequence as fixed effects and subject within sequence as a random effect. The inferential results (least-squares means [LSMs], difference between LSMs, and 90% CI of the difference) was exponentiated to the original scale. Geometric LSMs, geometric mean ratios (GMRs), and 90% CIs were presented.

### Cardiodynamic Data Analysis

#### QT-Interval Correction

The primary endpoint for evaluation of cardiodynamic effects in this study was (QTcF using the formula QT/(RR)1/3. If placebo-corrected change from baseline mean heart rate (ddHR) was >10 bpm (Garnett et al., 2018) during the drug evaluation days, population corrected (QTcP) was considered. Off-treatment data (predose and placebo) were used since the drug could change the QT-RR relationship. Mean ddHR > 10 bpm was examined (supported by the 90% CI of the mean ddHR) first using the following analysis of covariance (ANCOVA) model.

The effect of drug on HR was based on a linear mixed-effects model with the change from baseline dHR as dependent variable, dose (placebo, dose 1, dose 2, etc.), time (categorical), and dose by time as factors. Baseline values were used as a covariate. Subject was included in the model as random effect for the intercept with time point as the repeated variable. A 2-sided 90% CI was calculated for the contrast in active dose HR compared to placebo (CMS121 – placebo). Such a model was fitted across all nominal time points and was performed for the SAD and MAD parts separately.

### Statistical Analysis of Cardiodynamic Data

#### Categorical Analysis

Treatment-emergent counts were provided by treatment for the primary QTc correction categorized per the following ranges: ≤ 450, > 450 to ≤ 480, > 480 to ≤ 500 and > 500 msec, using the largest excursion in an individual subject once only. Counts were also provided by treatment and time point for QTc change from baseline values, categorized per the following ranges: < 30, ≥ 30 to ≤ 60, and > 60 msec. Categorical analysis was performed on QTcF (at a minimum) and on the QTc used for the exposure-response analysis, if different than QTcF.

#### Other ECG Parameters

Descriptive statistics including change from baseline were provided for QTc, QT, PR, and RR intervals, QRS duration, and HR by treatment and scheduled time point. The ddQTc (time-matched mean differences from placebo in dQTc) was also summarized by treatment and scheduled time point. Descriptive statistics for the cardiodynamic ECG parameters were generated using SAS^®^ Version 9.4.

## Results

### Study Population, Tolerability, and Safety

A total of 100 subjects (48 subjects in Part 1, 32 subjects in Part 2, 8 subjects in Part 3, and 12 subjects in Part 4) received at least 1 dose of study drug and were included in the safety analysis. The subject disposition, demographics and incidence of TEAEs in all subjects of the complete set of studies are summarized in Tables 2 and 3 (Supplemental).

**Table 2:**
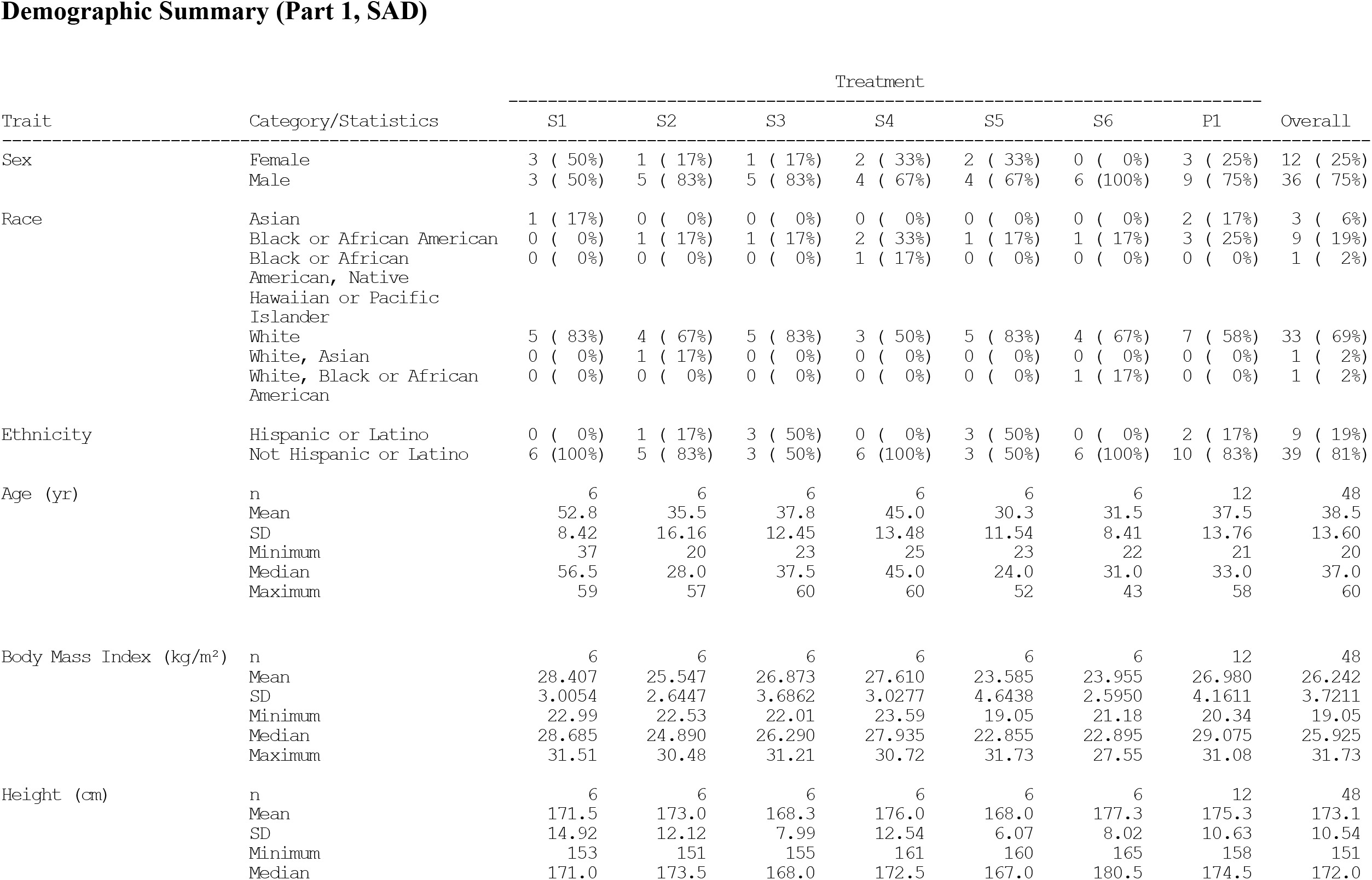
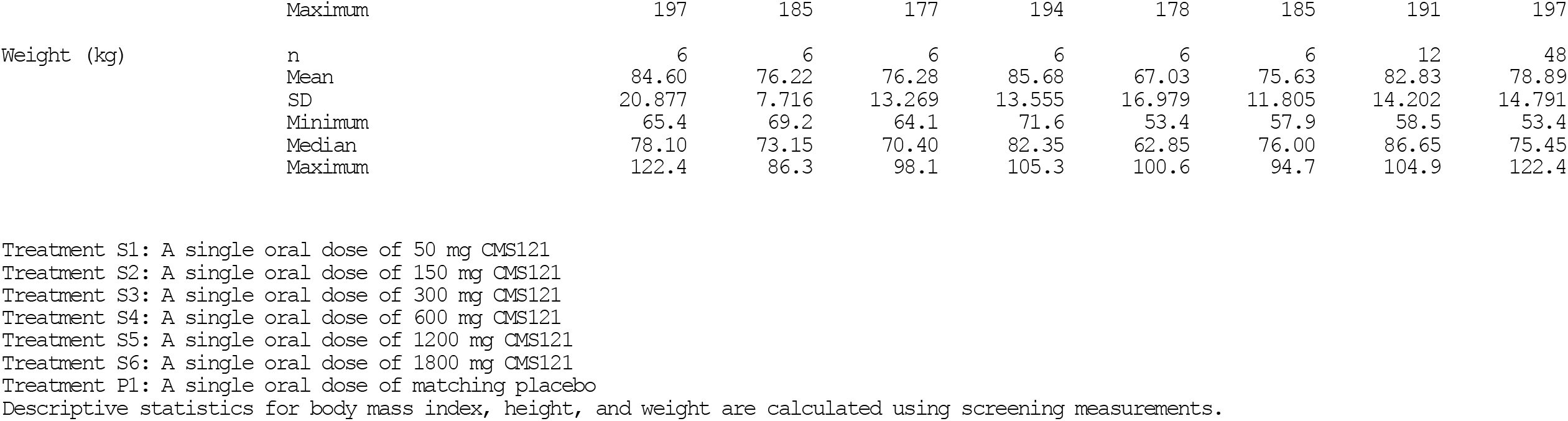
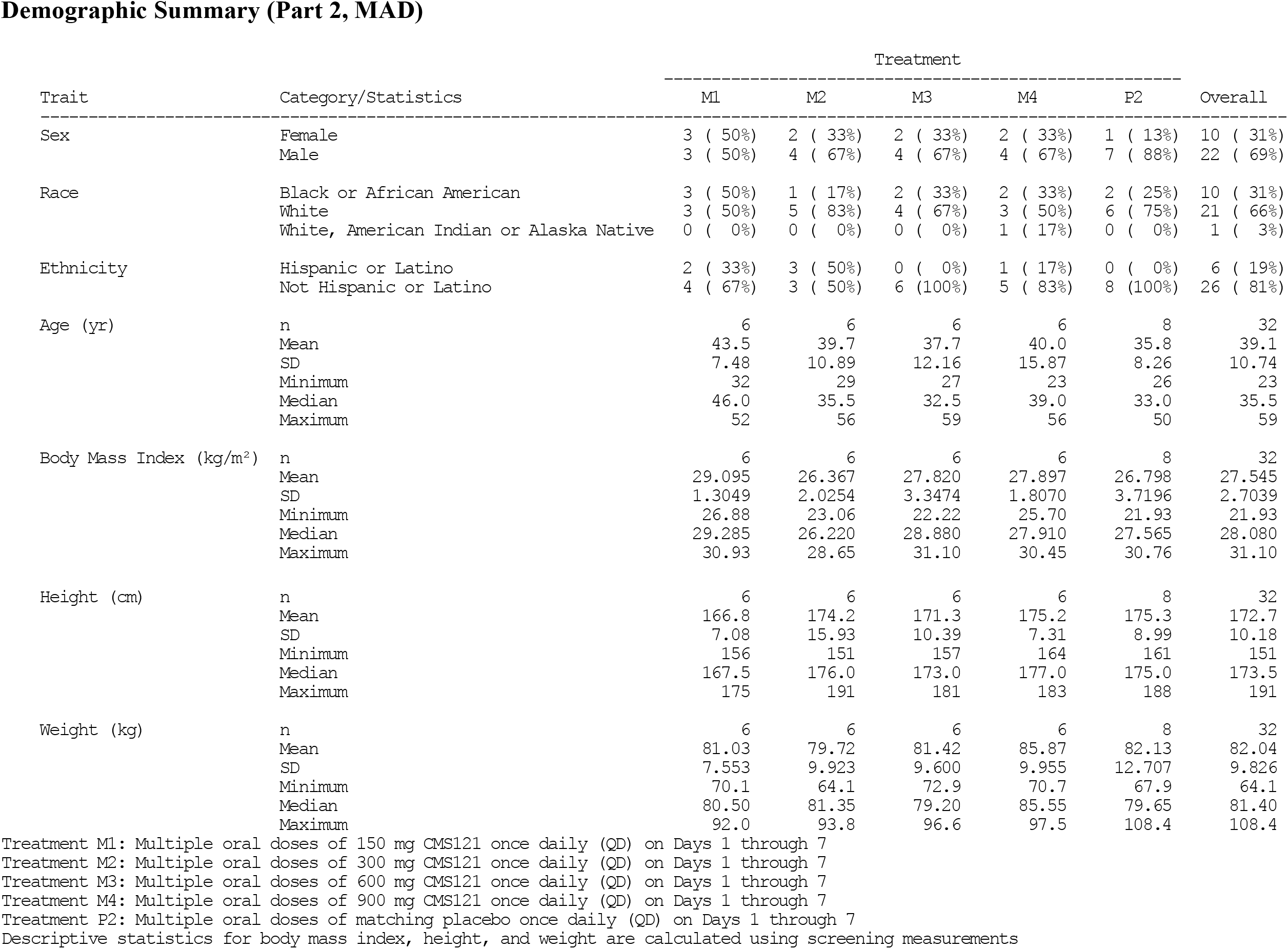
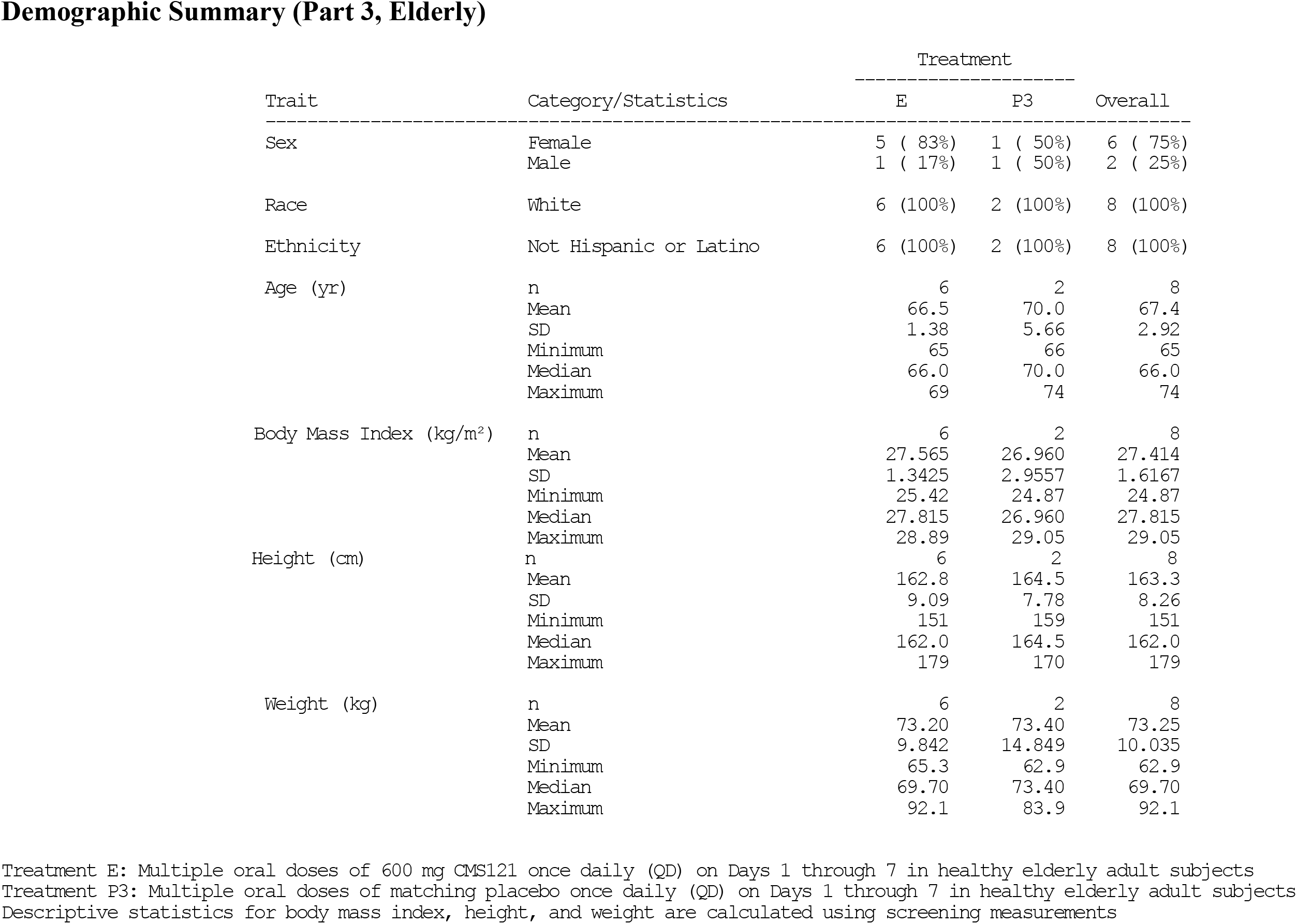
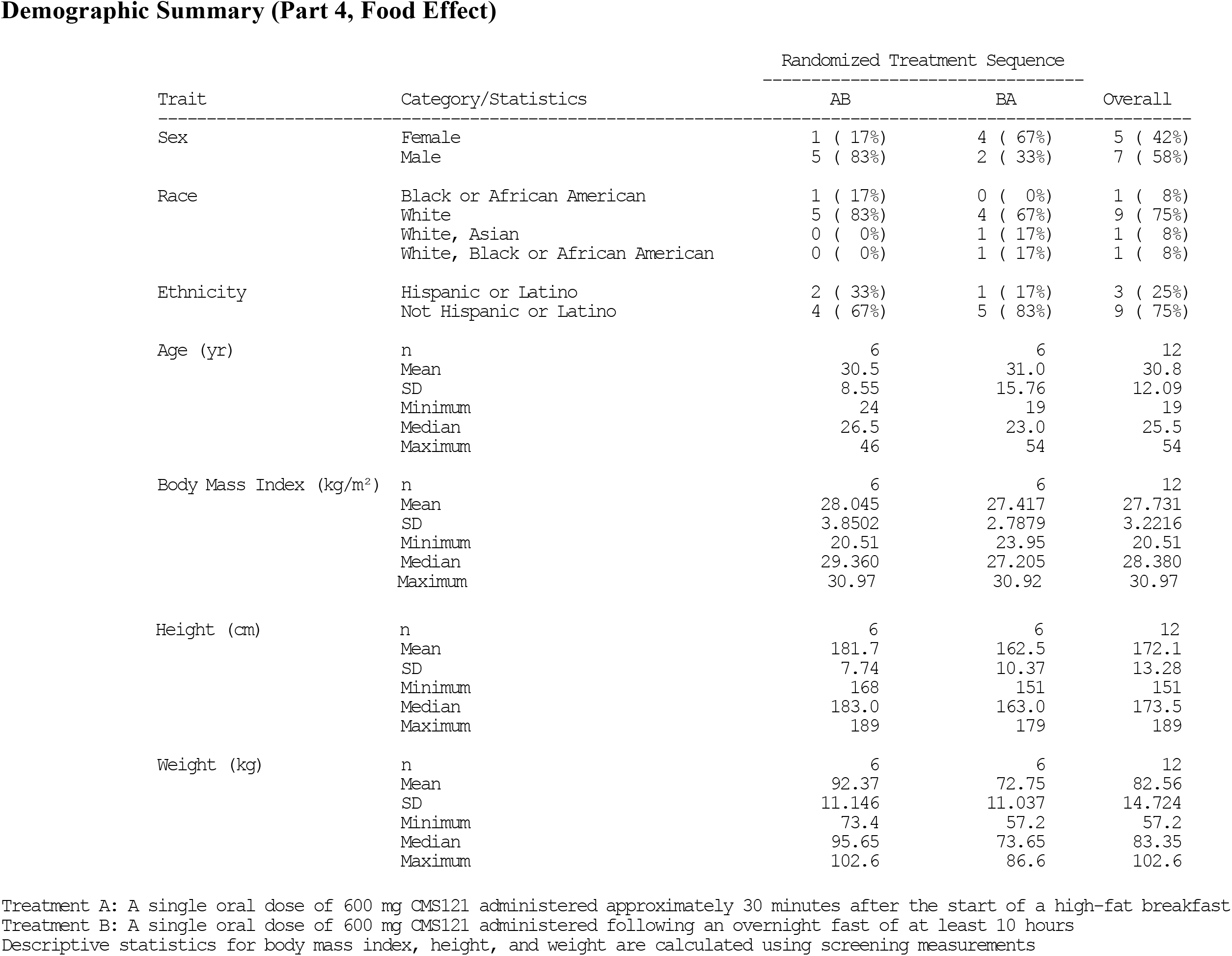
Subject baseline demographics of all subjects in the CMS121 Phase 1 series of Studies

**Table 3:**
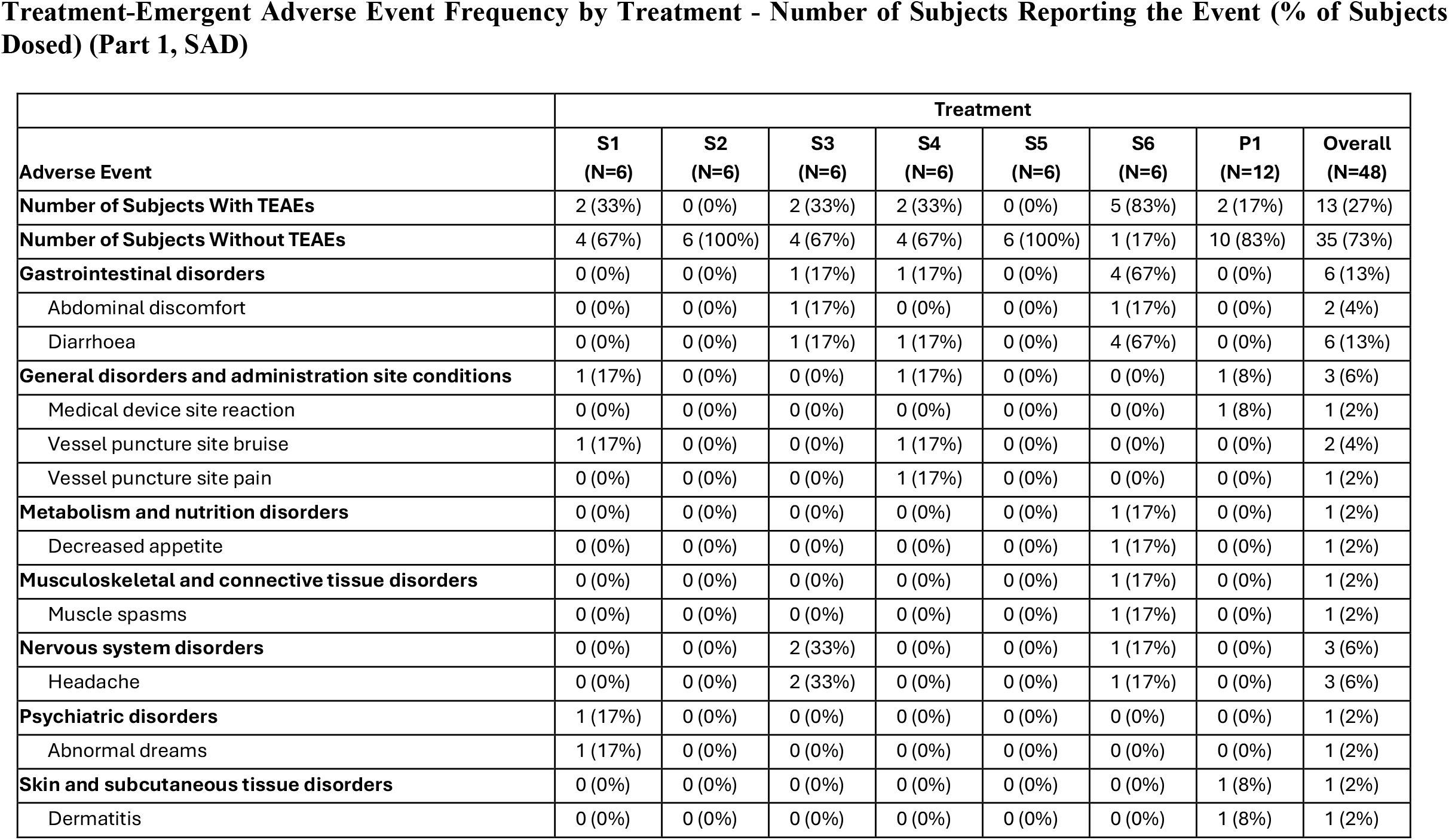
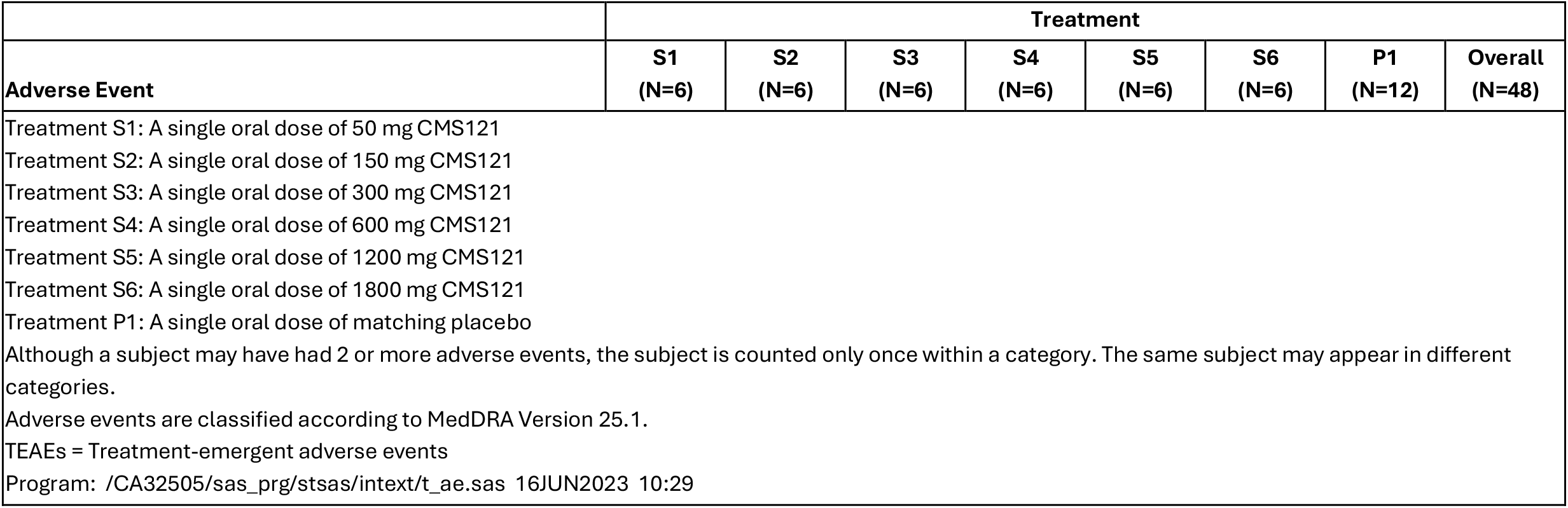
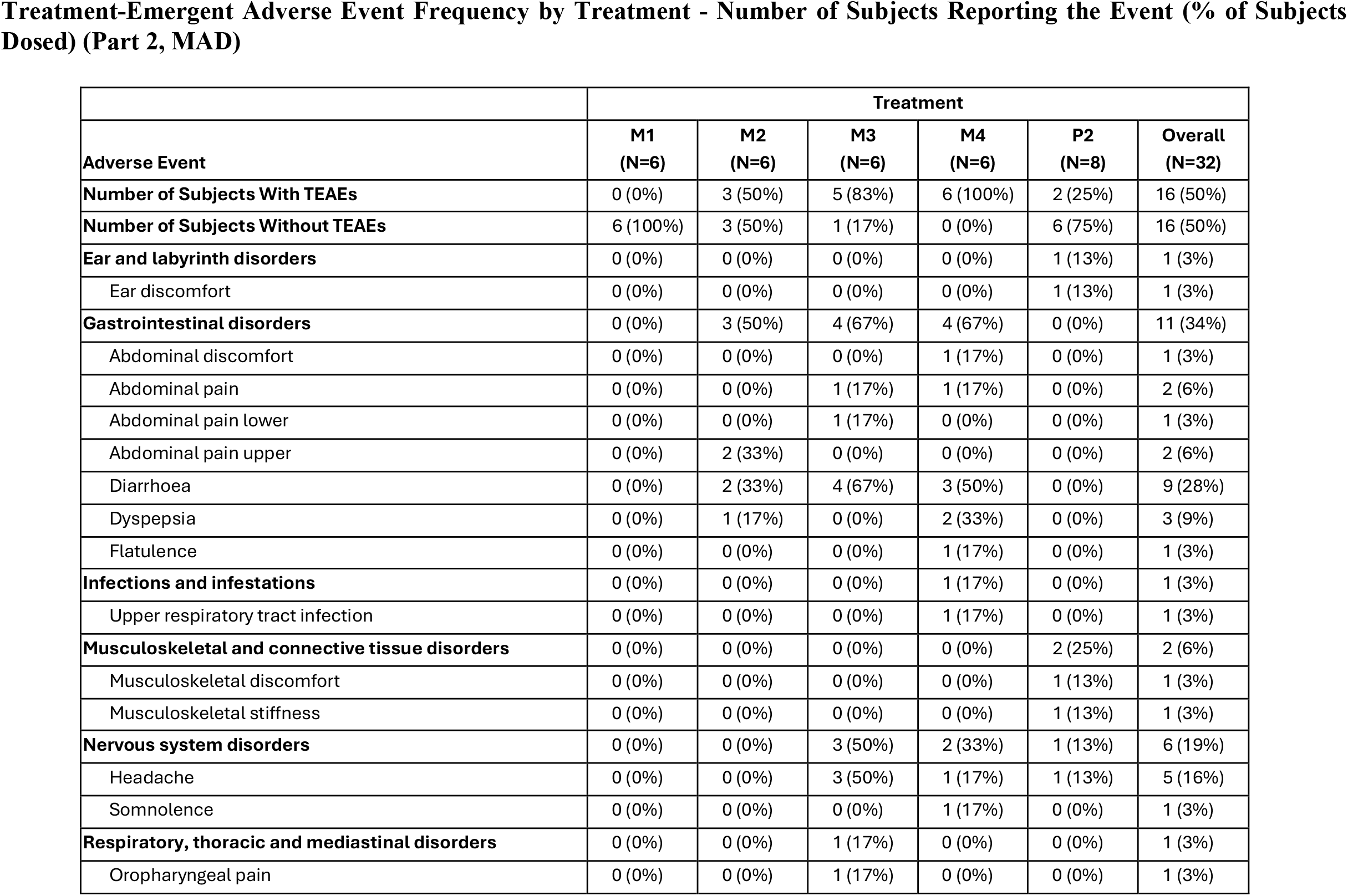
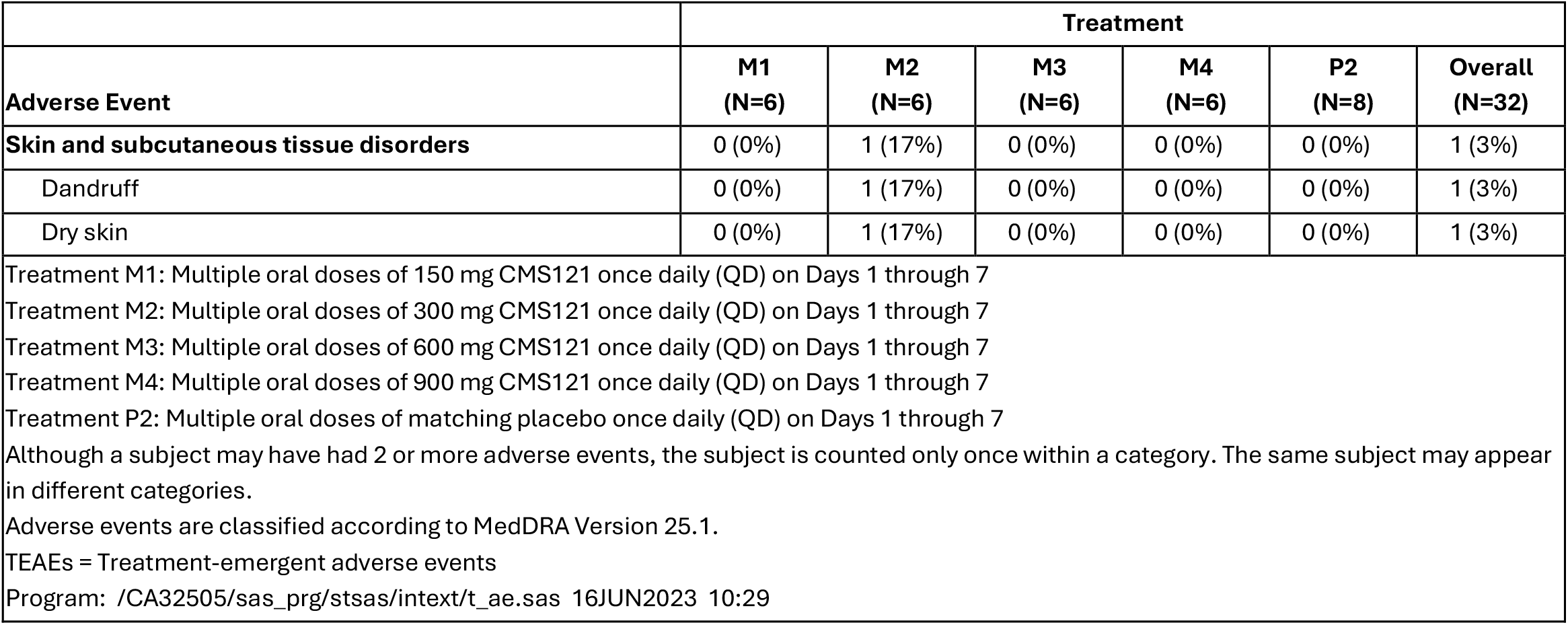
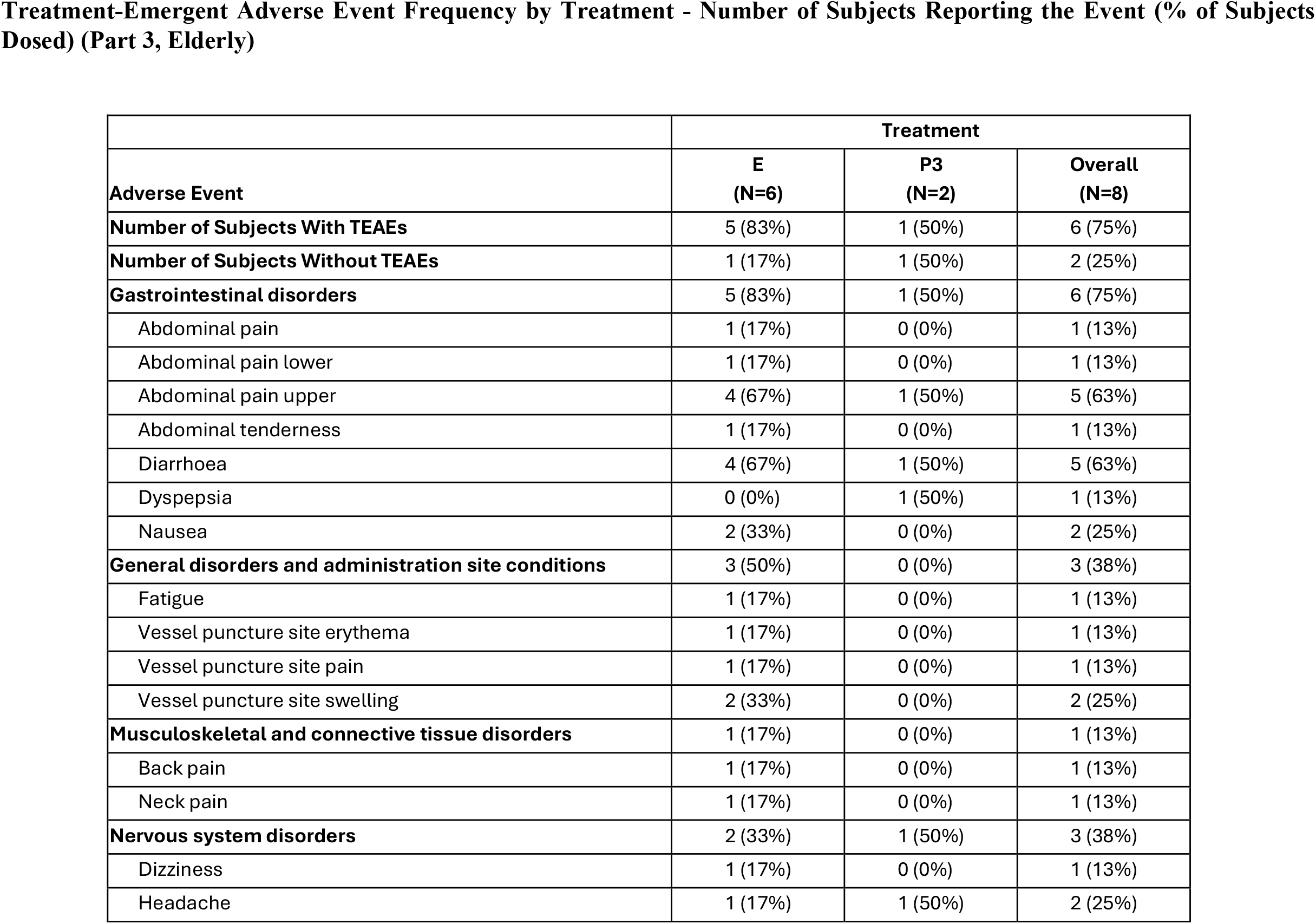
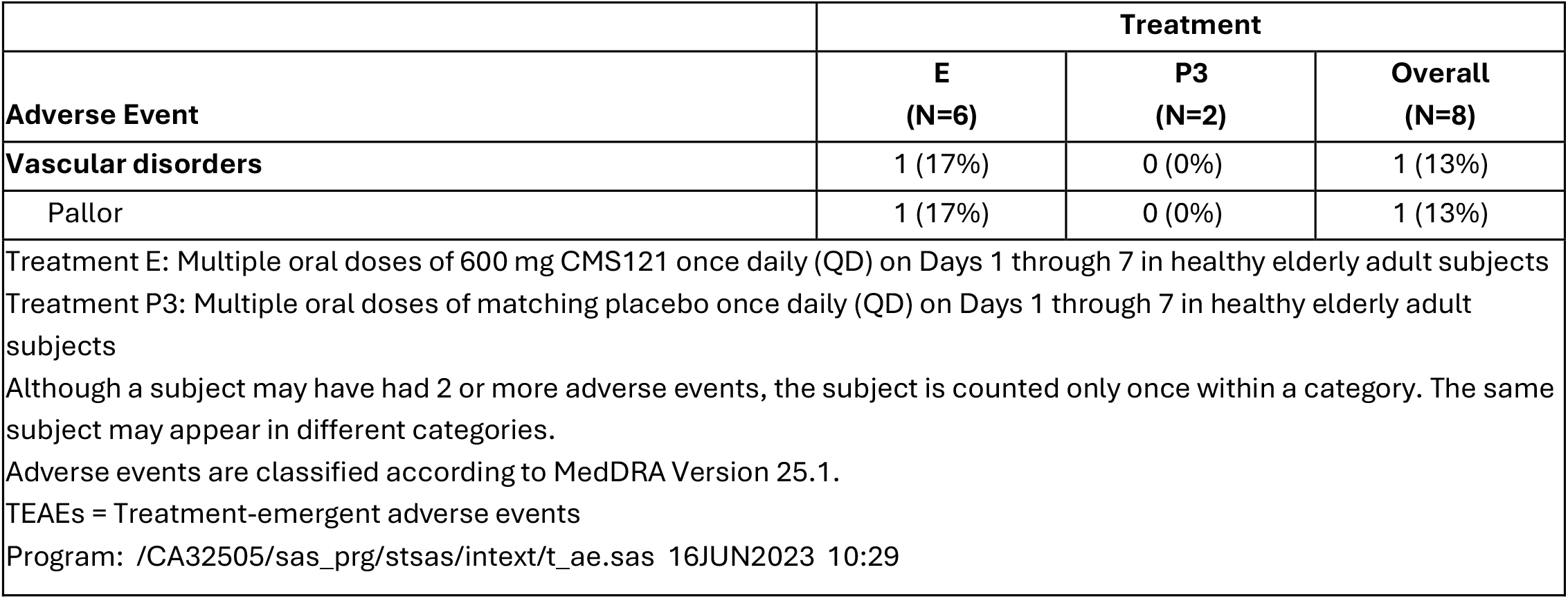
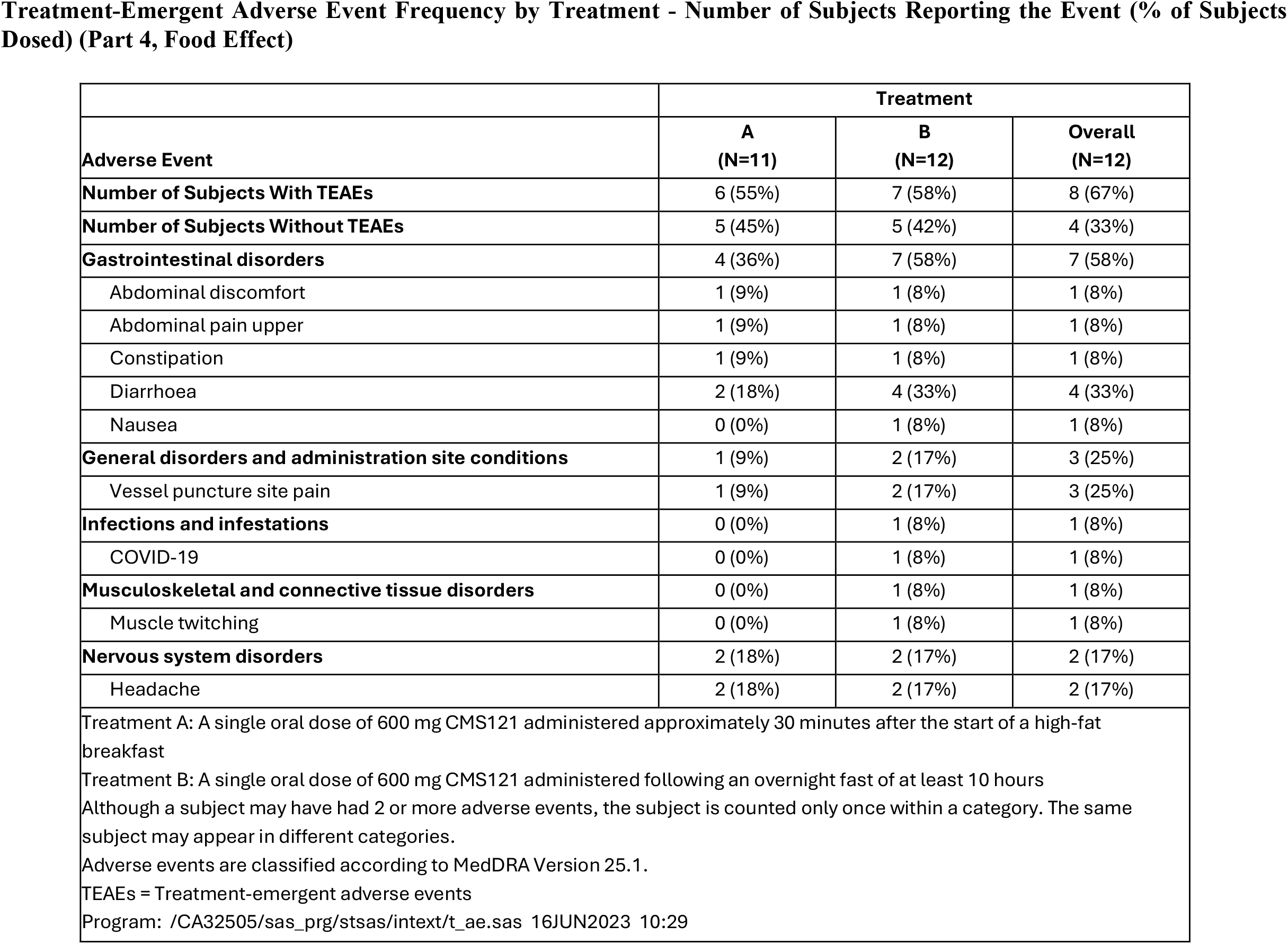
Incidence of TEAEs by subject and treatment in the CMS121 Phase 1 series of Studies

In Part 1, a total of 48 subjects (36 active treatment and 12 placebo) completed the study. In Part 2, a total of 31 subjects (24 active treatment and 7 placebo) completed all doses of study drug per protocol. One subject received only 2 doses of placebo before discontinuing from the study on Day 2 due to personal reasons. In Part 3, a total of 7 subjects (5 active treatment and 2 placebo) completed all doses of study drug. One subject received only 6 days of dosing with 600 mg CMS121 as dosing was held on Day 3 (due to an unrelated AE of dizziness). In Part 4, a total of 11 subjects completed all doses of study drug. One subject received only 1 dose of 600 mg CMS121 (fasted) before discontinuing from the study on Day 3 of Period 1 due to an AE of COVID-19. There were no deaths, no serious adverse events, and no severe treatment emergent adverse events (TEAEs). A total of 149 TEAEs were reported during the study. The majority (138; 93%) of the TEAEs were mild; 11 (7%) of the TEAEs were moderate. Those TEAEs which occurred in more than one subject and were more common in CMS121-treated subjects (compared to placebo-treated subjects) included gastrointestinal symptoms and headache. There were no clinically significant laboratory findings or laboratory AEs in the study. There were no clinically significant vital sign/physical examination findings or vital sign/physical examination AEs in the study. There were no abnormal postdose findings in the Columbia-Suicide Severity Rating Scale (C-SSRS). Safety electrocardiograms (ECGs) were analyzed; mean safety ECG parameters were within normal limits at the assessed post-dose time points and changes from baseline were small. Only one subject (single dose of 50 mg CMS121) had an isolated postdose (Day 1) QTcF that exceeded 450 msec (451 msec) and represented an increase from baseline of >30 msec (+32 msec). During the cardiodynamic portion of the study, there were no individual outliers with an absolute QTcF >450 msec or QTcF interval change from baseline ≥30 msec. CMS121 did not appear to prolong QTcF after single doses. However, after multiple doses of 300 and 600 mg CMS121 (not after 150 mg or 900 mg CMS121), statistical comparisons of dQTcF (between CMS121 and placebo) exceeded 10 msec at several time points, which could be an inconsistent, dose-independent effect of CMS121 (or its metabolites), an artifact of small sample size and/or an apparently lower baseline QTcF in the pooled placebo subjects.

### Pharmacokinetics

#### Part 1, SAD

Following administration of single oral doses of 50 mg to 1800 mg CMS121, CMS121 concentrations peaked at approximately 1.5 hours postdose, followed by biphasic elimination characterized by an initial rapid decline and a slower, sustained decline (Figures 2a and 2b). The concentration-time profiles were similar for the three metabolites, but CMS121-C2 accounted for the largest proportion of drug candidate in plasma, with its metabolite-to-parent ratio for Cmax and AUC0-inf exceeding 10 for all doses. As expected, CMS121 and its metabolites were better characterized at the higher doses as the AUC%extrap tended to decrease with increasing dose.

**Figure 2a.**
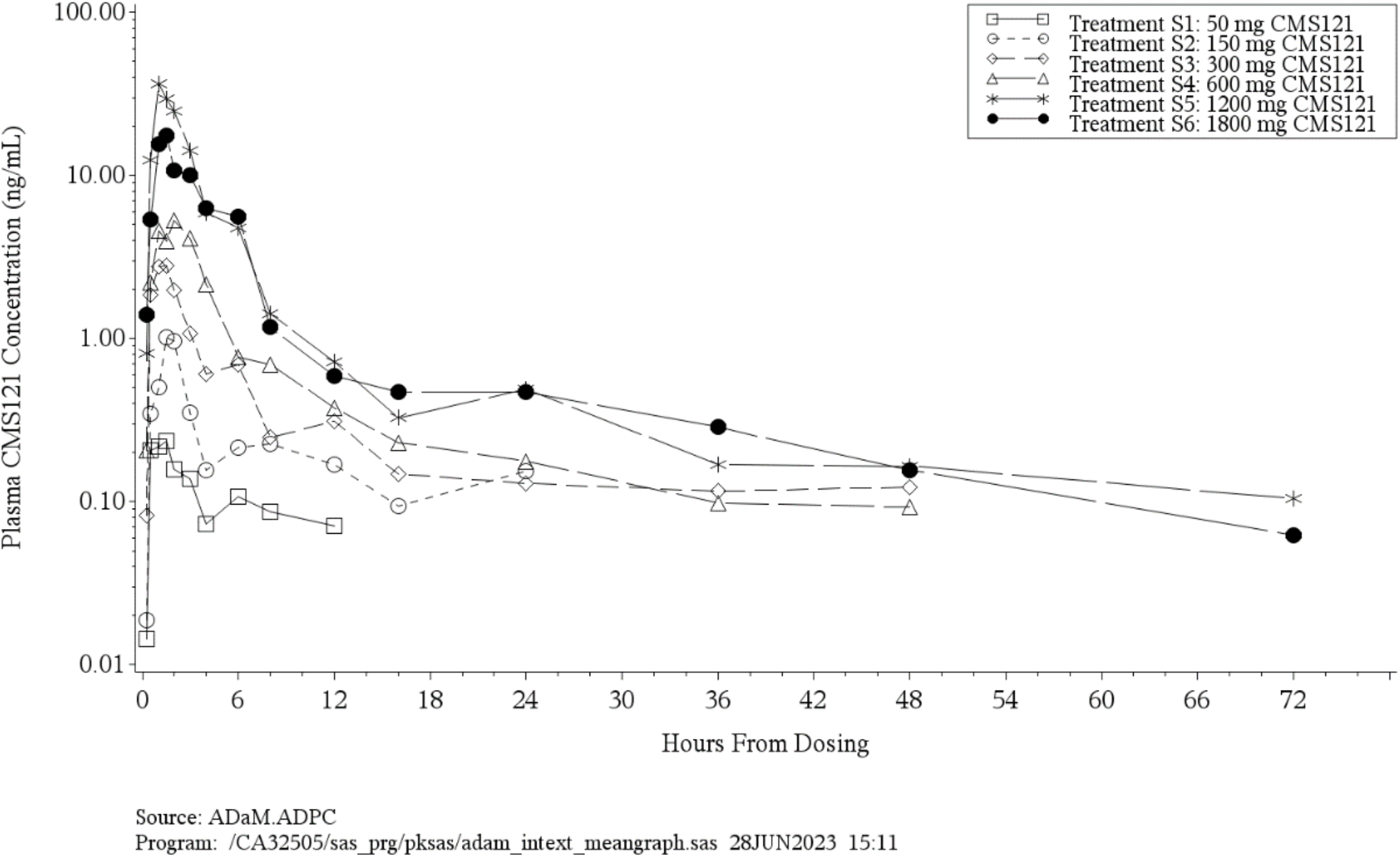
Arithmetic Mean <u>CMS121</u> Concentration Versus Time Profiles Following Single Oral Doses of 50 mg to 1800 mg CMS121 (Part 1, SAD) (Semi-Log Scale)

**Figure 2b.**
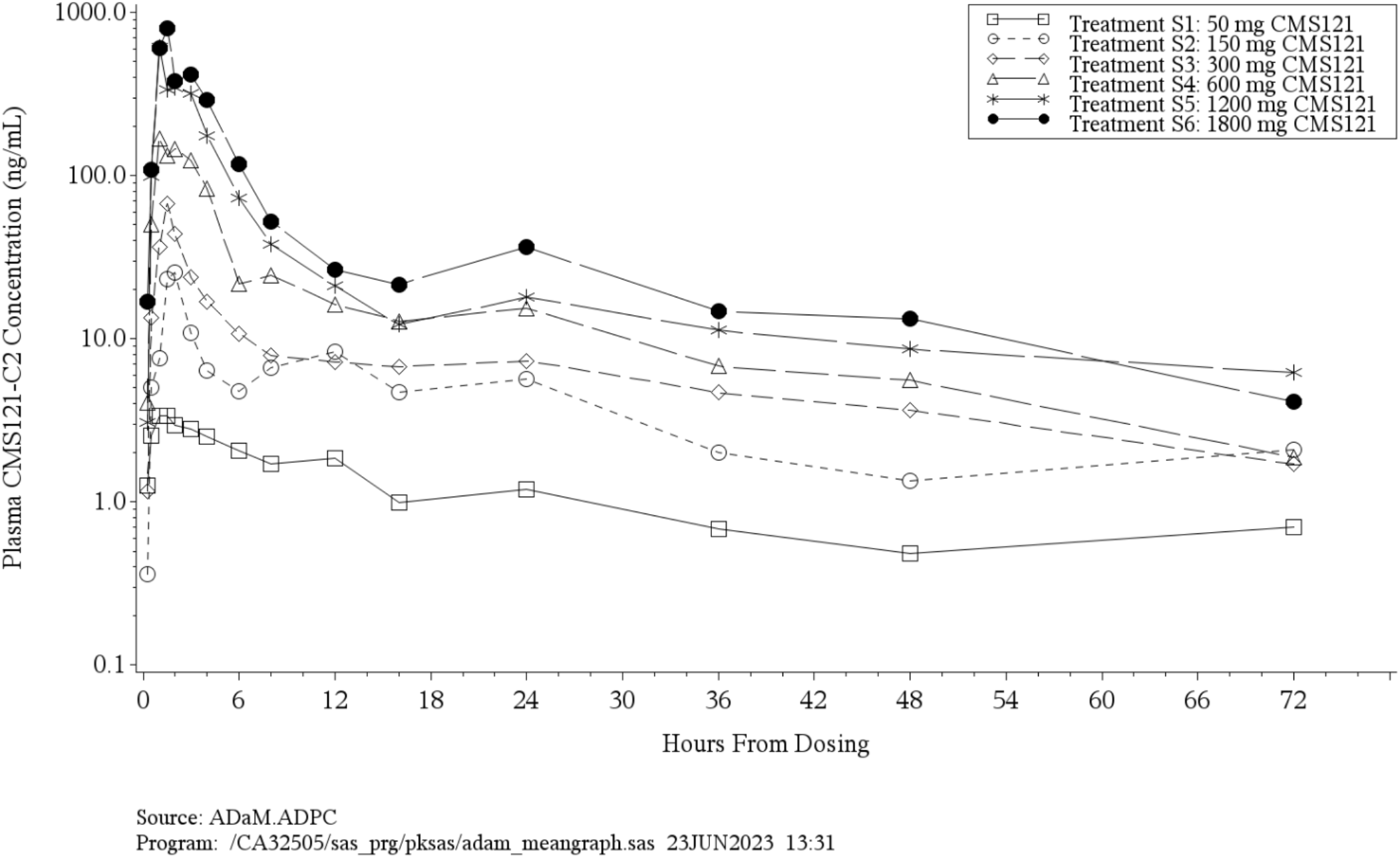
Arithmetic Mean <u>CMS121-C2</u> Concentration Versus Time Profiles Following Single Oral Doses of 50 mg to 1800 mg CMS121 (Part 1, SAD) (Semi-Log Scale)

When the SAD data was pooled with the single-dose data from the MAD cohorts, peak (Cmax) and overall (AUCs) exposure to CMS121 and CMS121-C1 displayed increases in a slightly greater than dose-proportional manner. For CMS121-C2, dose proportionality was shown for AUC0-inf, but Cmax, AUC0-t, and AUC0-24 were slightly greater than dose-proportional (Table 4a, 4b and 4c). Lastly, peak and overall exposure to CMS121-C3 increased in a dose-proportional manner, though this should be interpreted cautiously due to considerable variability in the data.

**Table 4a.** Summary of Plasma <u>CMS121</u> Pharmacokinetic Parameters Following Single Oral Doses of 50 mg to 1800 mg CMS121 (Part 1, SAD)

| Pharmacokinetic Parameters | Treatment S1 | Treatment S2 | Treatment S3 | Treatment S4 | Treatment S5 | Treatment S6 |
| --- | --- | --- | --- | --- | --- | --- |
| AUC<sub>0-t</sub> (ng h/mL) | 0.8306 (73.1) [n=6] | 3.602 (72.1) [n=6] | 10.37 (77.6) [n=6] | 23.08 (42.4) [n=6] | 94.94 (57.6) [n=6] | 69.74 (42.9) [n=6] |
| AUC<sub>0-24</sub> (ng h/mL) | 1.040 (82.0) [n=3] | 4.601 (41.5) [n=4] | 11.27 (68.1) [n=5] | 21.29 (45.0) [n=6] | 88.68 (58.2) [n=6] | 64.52 (39.5) [n=6] |
| AUC<sub>0-inf</sub> (ng h/mL) | 1.077 (87.7) [n=3] | 3.559 (45.0) [n=2] | 14.96 (62.5) [n=2] | 24.44 (39.9) [n=6] | 140.9 (72.1) [n=2] | 74.75 (45.0) [n=4] |
| AUC%extrap | 31.37 ± 24.251 [n=3] | 9.711 ± 3.9767 [n=2] | 7.351 ± 3.7468 [n=2] | 5.513 ± 3.7941 [n=6] | 1.201 ± 1.0931 [n=2] | 1.396 ± 0.74931 [n=4] |
| C<sub>max</sub> (ng/mL) | 0.3437 (88.0) [n=6] | 1.231 (100.7) [n=6] | 3.070 (110.6) [n=6] | 5.923 (81.5) [n=6] | 29.01 (118.9) [n=6] | 18.75 (54.6) [n=6] |
| T<sub>max</sub> (h) | 1.512 (0.50, 3.00) [n=6] | 1.501 (0.51, 6.00) [n=6] | 1.253 (1.00, 3.00) [n=6] | 1.752 (0.50, 4.00) [n=6] | 1.500 (0.50, 3.00) [n=6] | 1.517 (1.00, 3.00) [n=6] |
| T<sub>last</sub> (h) | 7.009 (4.00, 12.03) [n=6] | 14.013 (8.00, 24.01) [n=6] | 24.039 (8.01, 48.00) [n=6] | 36.001 (12.00, 48.00) [n=6] | 60.039 (24.00, 72.03) [n=6] | 48.000 (24.04, 72.00) [n=6] |
| K<sub>el</sub> (1/h) | 0.3948 ± 0.43451 [n=3] | 0.2545 ± 0.013381 [n=2] | 0.09269 ± 0.0082244 [n=2] | 0.1244 ± 0.15050 [n=6] | 0.1153 ± 0.12031 [n=2] | 0.1051 ± 0.061804 [n=4] |
| t<sub>1/2</sub> (h) | 3.637 ± 2.7477 [n=3] | 2.728 ± 0.1434 [n=2] | 7.507 ± 0.6661 [n=2] | 9.596 ± 4.1908 [n=6] | 13.179 ± 13.7472 [n=2] | 8.264 ± 3.9746 [n=4] |
| CL/F (L/h) | 55240 ± 36645 [n=3] | 44110 ± 18389 [n=2] | 21730 ± 11835 [n=2] | 26160 ± 10451 [n=6] | 9426 ± 5707.9 [n=2] | 25580 ± 8926.7 [n=4] |
| V<sub>z</sub>/F (L) | 257000 ± 241640 [n=3] | 171700 ± 63241 [n=2] | 241000 ± 149070 [n=2] | 368100 ± 217240 [n=6] | 122600 ± 78412 [n=2] | 299400 ± 204530 [n=4] |
| Treatment S1: A single oral dose of 50 mg CMS121<br>Treatment S2: A single oral dose of 150 mg CMS121<br>Treatment S3: A single oral dose of 300 mg CMS121<br>Treatment S4: A single oral dose of 600 mg CMS121<br>Treatment S5: A single oral dose of 1200 mg CMS121<br>Treatment S6: A single oral dose of 1800 mg CMS121<br>AUCs and Cmax are presented as geometric mean and geometric CV%.<br>Tmax and Tlast are presented as median (minimum, maximum).<br>Other parameters are presented as arithmetic mean ± SD.<br>Program: /CA32505/sas_prg/pksas/adam_intext_stats_steady_state.sas 15JUN2023 16:08 |  |  |  |  |  |  |

**Table 4b.** Summary of Plasma <u>CMS121-C2</u> Pharmacokinetic Parameters Following Single Oral Doses of 50 mg to 1800 mg CMS121 (Part 1, SAD)

| Pharmacokinetic Parameters | Treatment S1 | Treatment S2 | Treatment S3 | Treatment S4 | Treatment S5 | Treatment S6 |
| --- | --- | --- | --- | --- | --- | --- |
| AUC0-t (ng h/mL) | 56.15 (36.8) [n=6] | 210.8 (75.9) [n=6] | 318.9 (110.5) [n=6] | 1024 (56.6) [n=6] | 2229 (34.5) [n=6] | 3129 (45.1) [n=6] |
| AUC0-24 (ng h/mL) | 39.04 (34.6) [n=6] | 147.9 (70.4) [n=6] | 220.6 (87.5) [n=6] | 780.9 (47.2) [n=6] | 1805 (28.3) [n=6] | 2521 (41.9) [n=6] |
| AUC0-inf (ng h/mL) | 76.15 (33.9) [n=5] | 347.7 (37.0) [n=4] | 529.3 (73.0) [n=4] | 1186 (71.1) [n=4] | 2235 (49.2) [n=4] | 3262 (43.4) [n=6] |
| AUC%extrap | 22.12 ± 13.408 [n=5] | 10.67 ± 10.507 [n=4] | 5.450 ± 4.2241 [n=4] | 2.853 ± 1.8871 [n=4] | 6.680 ± 6.5919 [n=4] | 4.011 ± 3.9676 [n=6] |
| Cmax (ng/mL) | 5.305 (62.5) [n=6] | 23.46 (103.3) [n=6] | 44.77 (132.3) [n=6] | 190.3 (94.8) [n=6] | 622.2 (83.7) [n=6] | 838.8 (83.6) [n=6] |
| Tmax (h) | 2.288 (0.50, 4.00) [n=6] | 1.750 (0.51, 3.00) [n=6] | 1.002 (1.00, 3.00) [n=6] | 2.002 (1.05, 4.00) [n=6] | 1.999 (1.00, 3.00) [n=6] | 2.501 (1.00, 4.02) [n=6] |
| Tlast (h) | 48.008 (24.07, 72.01) [n=6] | 48.008 (48.00, 72.04) [n=6] | 48.006 (36.01, 72.03) [n=6] | 72.004 (48.00, 72.02) [n=6] | 72.002 (48.11, 72.03) [n=6] | 72.000 (48.00, 72.04) [n=6] |
| Kel (1/h) | 0.04850 ± 0.035191 [n=5] | 0.06581 ± 0.042060 [n=4] | 0.06682 ± 0.026186 [n=4] | 0.04491 ± 0.0095639 [n=4] | 0.04625 ± 0.035587 [n=4] | 0.05073 ± 0.035091 [n=6] |
| t½ (h) | 22.493 ± 15.0560 [n=5] | 18.165 ± 18.0402 [n=4] | 11.692 ± 4.7467 [n=4] | 16.094 ± 4.1599 [n=4] | 21.905 ± 13.2739 [n=4] | 20.503 ± 12.7337 [n=6] |
| MR AUC0-inf | 44.740 ± 41.0838 [n=2] | 55.259 ± . [n=1] | 19.939 ± 8.9026 [n=2] | 42.040 ± 25.7236 [n=4] | 10.104 ± . [n=1] | 28.556 ± 10.9449 [n=4] |
| MR Cmax | 10.616 ± 4.3855 [n=6] | 13.531 ± 6.3418 [n=6] | 11.411 ± 7.9226 [n=6] | 28.409 ± 24.9935 [n=6] | 16.004 ± 9.2122 [n=6] | 31.378 ± 13.4014 [n=6] |
| Treatment S1: A single oral dose of 50 mg CMS121 |  |  |  |  |  |  |
| Treatment S2: A single oral dose of 150 mg CMS121 |  |  |  |  |  |  |
| Treatment S3: A single oral dose of 300 mg CMS121 |  |  |  |  |  |  |
| Treatment S4: A single oral dose of 600 mg CMS121 |  |  |  |  |  |  |
| Treatment S5: A single oral dose of 1200 mg CMS121 |  |  |  |  |  |  |
| Treatment S6: A single oral dose of 1800 mg CMS121 |  |  |  |  |  |  |
| AUCs and Cmax are presented as geometric mean and geometric CV%. |  |  |  |  |  |  |
| Tmax and Tlast are presented as median (minimum, maximum). |  |  |  |  |  |  |
| Other parameters are presented as arithmetic mean ± SD. |  |  |  |  |  |  |
| Program: /CA32505/sas_prg/pksas/adam_intext_stats_steady_state.sas 15JUN2023 16:08 |  |  |  |  |  |  |

**Table 4c.**
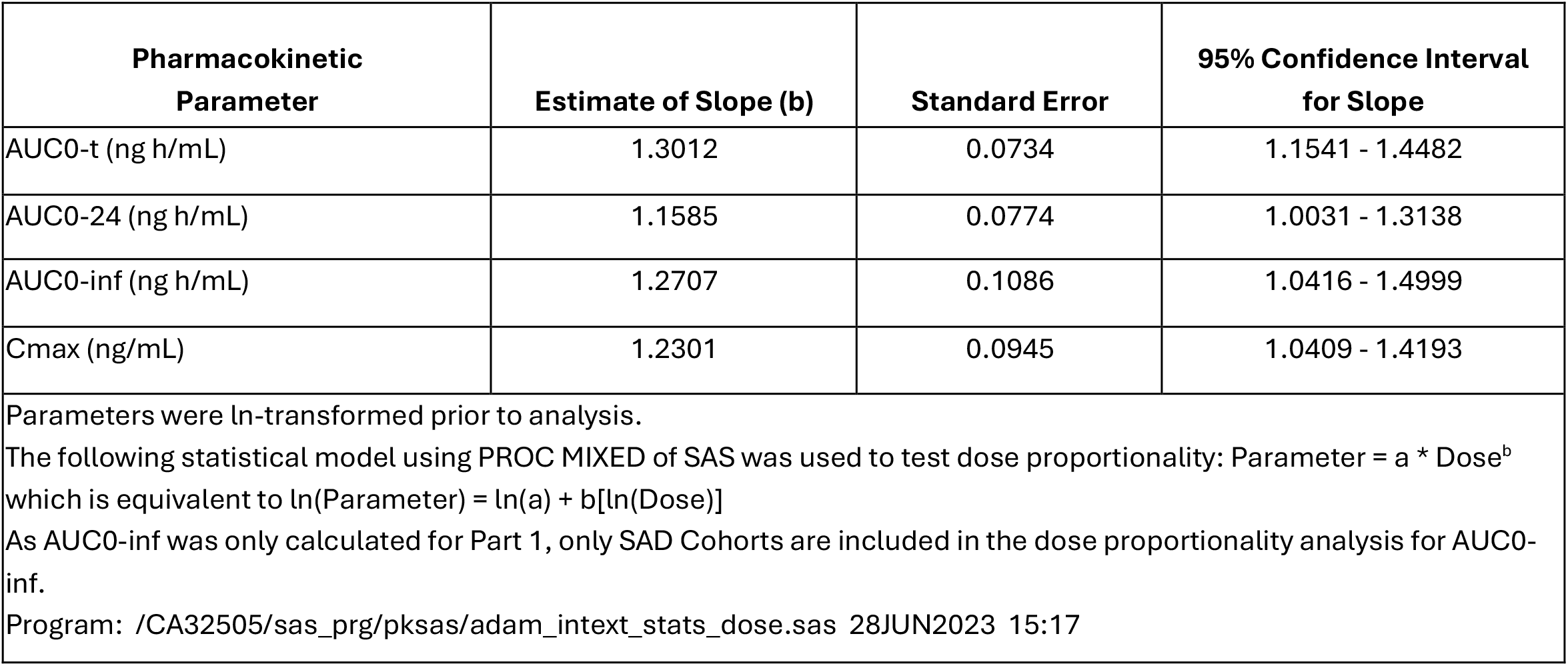
Summary of the Dose Proportionality Analysis of Plasma CMS121 Pharmacokinetic Parameters Following Single Oral Doses of 50 mg to 1800 mg CMS121 (Part 1, SAD, and Part 2, MAD, Day 1)

| Pharmacokinetic Parameter | Estimate of Slope (b) | Standard Error | 95% Confidence Interval for Slope |
| --- | --- | --- | --- |
| AUC0-t (ng h/mL) | 1.3012 | 0.0734 | 1.1541 - 1.4482 |
| AUC0-24 (ng h/mL) | 1.1585 | 0.0774 | 1.0031 - 1.3138 |
| AUC0-inf (ng h/mL) | 1.2707 | 0.1086 | 1.0416 - 1.4999 |
| Cmax (ng/mL) | 1.2301 | 0.0945 | 1.0409 - 1.4193 |
| Parameters were ln-transformed prior to analysis.<br>The following statistical model using PROC MIXED of SAS was used to test dose proportionality: $\text{Parameter} = a * \text{Dose}^b$ which is equivalent to $\ln(\text{Parameter}) = \ln(a) + b[\ln(\text{Dose})]$<br>As AUC0-inf was only calculated for Part 1, only SAD Cohorts are included in the dose proportionality analysis for AUC0-inf.<br>Program: /CA32505/sas_prg/pksas/adam_intext_stats_dose.sas 28JUN2023 15:17 |  |  |  |

Urinary excretion of CMS121 metabolites was minimal, with the mean cumulative fraction excreted at 72 hours postdose being 0.226% or less for all doses.

#### Part 2, MAD

On Day 7, following the administration of multiple oral doses of 150 mg to 900 mg CMS121 QD, CMS121 concentrations peaked at approximately 2-3 hours postdose, followed by a similar biphasic elimination that was observed for the SAD cohorts (Figures 3a and 3b). The concentration-time profiles were similar for the three metabolites, with CMS121-C2 also accounting for the largest proportion of drug in plasma, with its metabolite-to-parent ratio for Cmax,ss and AUCtau exceeding 10 for most doses (Table 5a and 5b).

**Table 5a.**
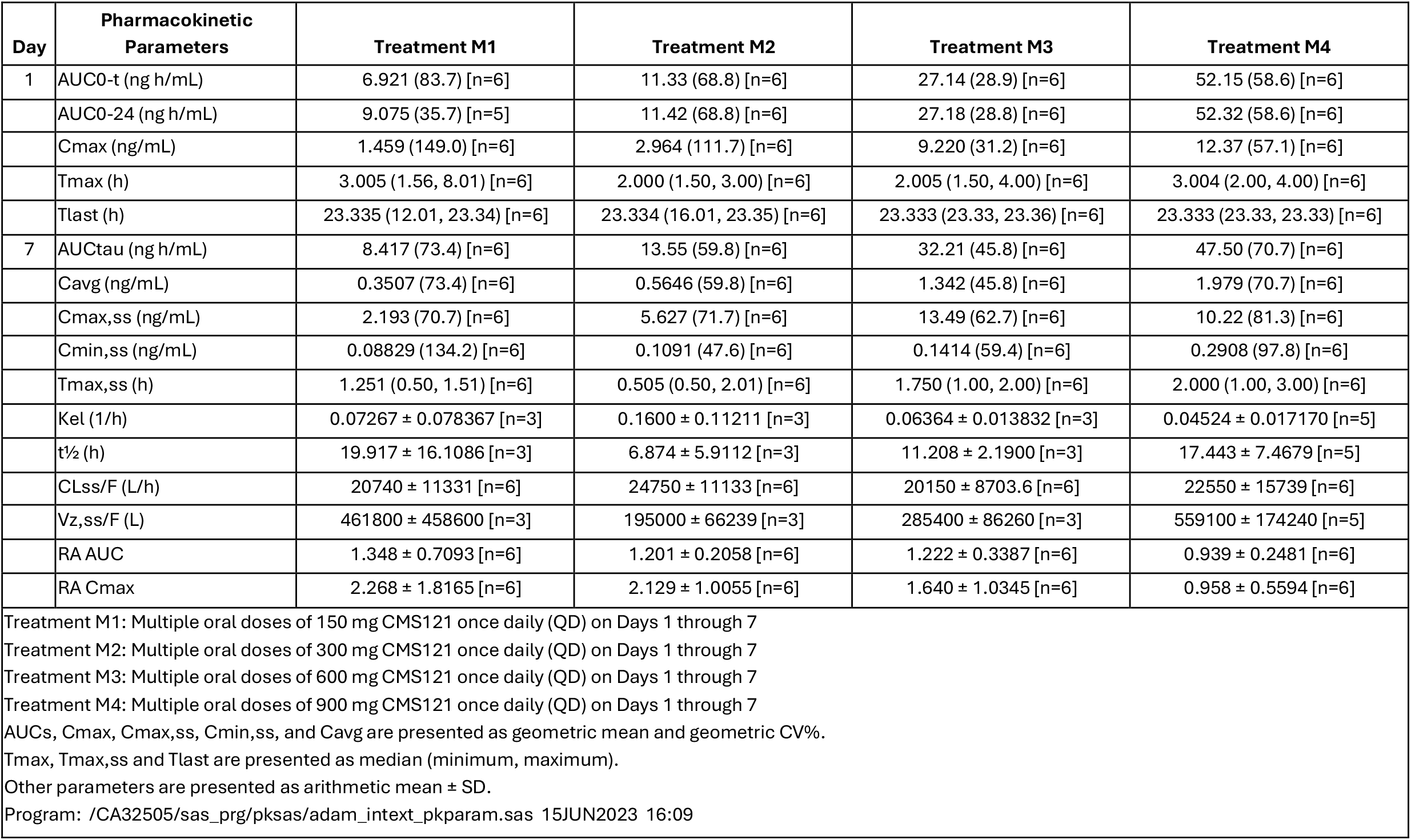
Summary of Plasma <u>CMS121</u> Pharmacokinetic Parameters Following Multiple Oral Doses of 150 mg to 900 mg CMS121 QD (**Part 2, MAD)**

**Table 5b.** Summary of Plasma <u>CMS121-C2</u> Pharmacokinetic Parameters Following Multiple Oral Doses of 150 mg to 900 mg CMS121 QD **(Part 2, MAD)**

| Day | Pharmacokinetic Parameters | Treatment M1 | Treatment M2 | Treatment M3 | Treatment M4 |
| --- | --- | --- | --- | --- | --- |
| 1 | AUC<sub>0-t</sub> (ng h/mL) | 209.5 (61.4) [n=6] | 341.0 (27.4) [n=6] | 675.0 (57.8) [n=6] | 1686 (47.6) [n=6] |
|  | AUC<sub>0-24</sub> (ng h/mL) | 210.2 (61.0) [n=6] | 341.7 (27.3) [n=6] | 675.2 (57.9) [n=6] | 1692 (47.3) [n=6] |
|  | C<sub>max</sub> (ng/mL) | 23.32 (107.2) [n=6] | 61.71 (90.3) [n=6] | 242.5 (71.5) [n=6] | 433.0 (36.5) [n=6] |
|  | T<sub>max</sub> (h) | 5.003 (1.56, 23.33) [n=6] | 2.001 (1.99, 3.00) [n=6] | 2.500 (1.50, 4.00) [n=6] | 3.004 (2.00, 4.00) [n=6] |
|  | T<sub>last</sub> (h) | 23.335 (23.33, 23.34) [n=6] | 23.335 (23.33, 23.35) [n=6] | 23.333 (23.33, 23.36) [n=6] | 23.333 (23.33, 23.33) [n=6] |
| 7 | AUC<sub>tau</sub> (ng h/mL) | 308.0 (80.1) [n=6] | 349.7 (39.2) [n=6] | 864.1 (61.8) [n=6] | 1416 (39.3) [n=6] |
|  | C<sub>avg</sub> (ng/mL) | 12.83 (80.1) [n=6] | 14.57 (39.2) [n=6] | 36.01 (61.8) [n=6] | 58.98 (39.3) [n=6] |
|  | C<sub>max,ss</sub> (ng/mL) | 42.39 (89.6) [n=6] | 78.02 (70.8) [n=6] | 290.8 (82.2) [n=6] | 367.6 (51.4) [n=6] |
|  | C<sub>min,ss</sub> (ng/mL) | 5.563 (151.1) [n=6] | 4.937 (98.3) [n=6] | 7.657 (60.7) [n=6] | 12.12 (81.7) [n=6] |
|  | T<sub>max,ss</sub> (h) | 1.503 (0.50, 4.00) [n=6] | 1.038 (1.00, 2.01) [n=6] | 2.000 (1.00, 2.00) [n=6] | 2.000 (0.51, 3.00) [n=6] |
|  | K<sub>el</sub> (1/h) | 0.03239 ± 0.018634 [n=5] | 0.05439 ± 0.022826 [n=5] | 0.06240 ± 0.046558 [n=5] | 0.04925 ± 0.023039 [n=6] |
|  | t<sub>1/2</sub> (h) | 25.409 ± 9.0543 [n=5] | 14.478 ± 5.4212 [n=5] | 17.177 ± 12.3703 [n=5] | 18.957 ± 12.8499 [n=6] |
|  | RA AUC | 1.648 ± 0.7922 [n=6] | 1.046 ± 0.2319 [n=6] | 1.313 ± 0.2974 [n=6] | 0.871 ± 0.2585 [n=6] |
|  | RA C<sub>max</sub> | 2.182 ± 1.2674 [n=6] | 1.577 ± 1.1173 [n=6] | 1.364 ± 0.7912 [n=6] | 1.000 ± 0.5613 [n=6] |
|  | MR AUC<sub>tau</sub> | 25.754 ± 10.9345 [n=6] | 17.148 ± 4.0192 [n=6] | 18.149 ± 6.6527 [n=6] | 21.540 ± 12.0032 [n=6] |
|  | MR C<sub>max,ss</sub> | 13.850 ± 6.2683 [n=6] | 9.788 ± 4.1648 [n=6] | 15.458 ± 8.0253 [n=6] | 25.963 ± 14.6619 [n=6] |
| Treatment M1: Multiple oral doses of 150 mg CMS121 once daily (QD) on Days 1 through 7<br>Treatment M2: Multiple oral doses of 300 mg CMS121 once daily (QD) on Days 1 through 7<br>Treatment M3: Multiple oral doses of 600 mg CMS121 once daily (QD) on Days 1 through 7<br>Treatment M4: Multiple oral doses of 900 mg CMS121 once daily (QD) on Days 1 through 7<br>AUCs, C<sub>max</sub>, C<sub>max,ss</sub>, C<sub>min,ss</sub>, and C<sub>avg</sub> are presented as geometric mean and geometric CV%.<br>T<sub>max</sub>, T<sub>max,ss</sub> and T<sub>last</sub> are presented as median (minimum, maximum).<br>Other parameters are presented as arithmetic mean ± SD.<br>Program: /CA32505/sas_prg/pksas/adam_intext_pkparam.sas 15JUN2023 16:09 |  |  |  |  |  |

**Figure 3a.**
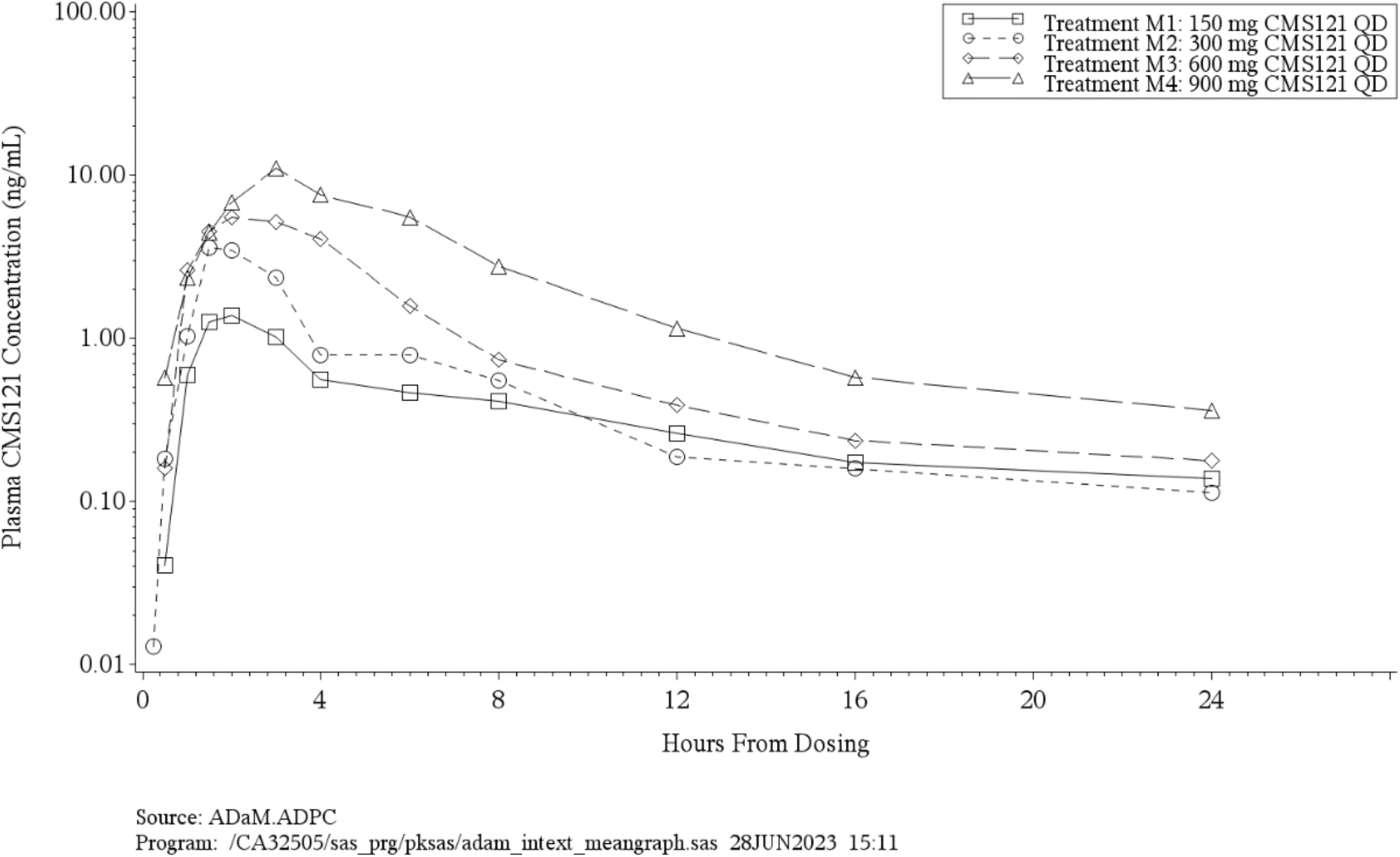
Arithmetic Mean <u>CMS121</u> Concentration Versus Time Profiles Following the First Oral Dose of 150 mg to 900 mg CMS121 (Part 2, <u>MAD, Day 1</u>) (Semi-Log Scale)

**Figure 3b.**
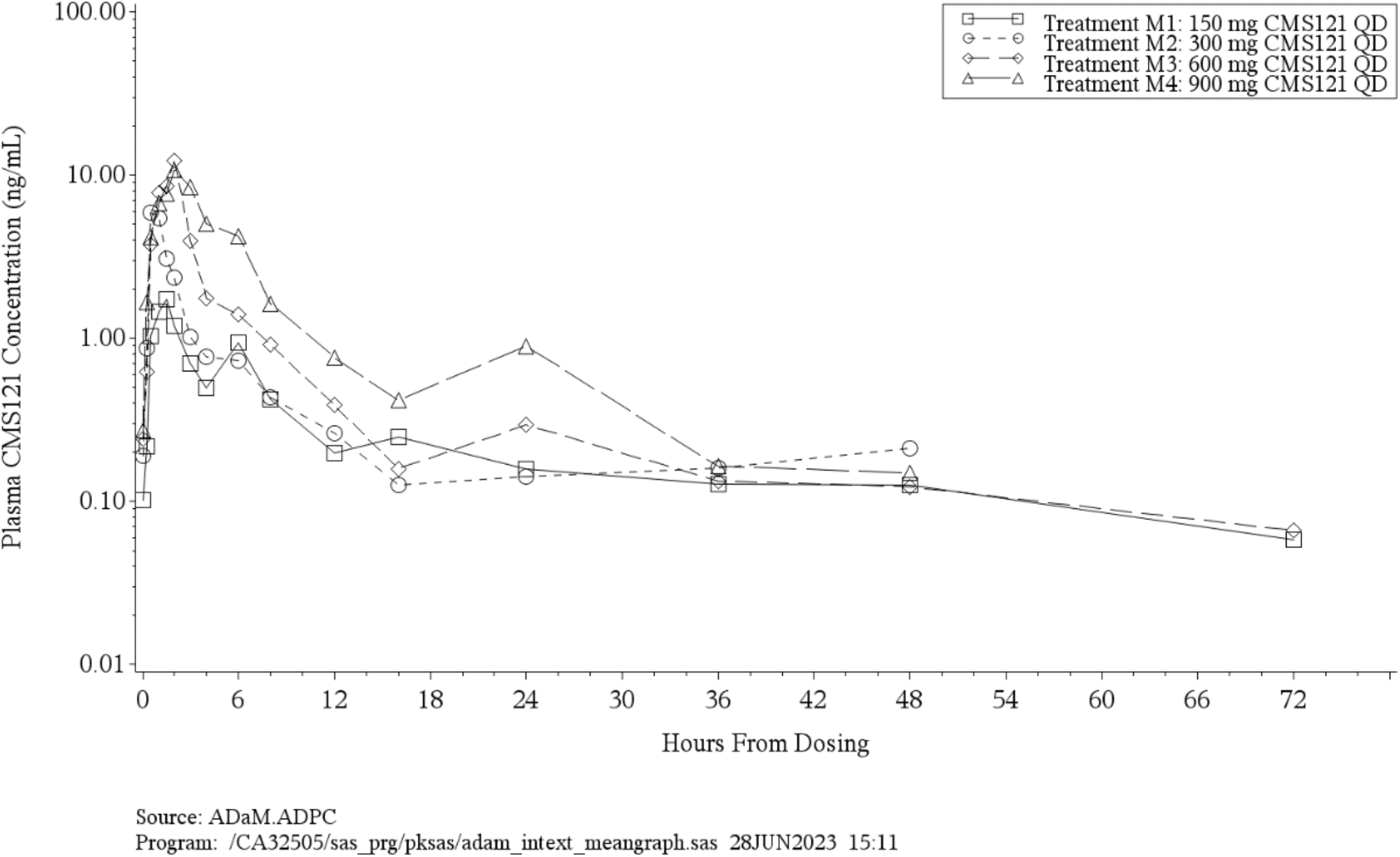
Arithmetic Mean <u>CMS121</u> Concentration Versus Time Profiles Following Multiple Oral Doses of 150 mg to 900 mg CMS121 QD (<u>Part 2, MAD, Day 7</u>) (Semi-Log Scale)

CMS121 and its metabolites did not display any appreciable accumulation in total exposure over time but peak concentrations did accumulate, especially at the lower doses, for the parent, CMS121-C1, and CMS121-C2. Negligible accumulation was observed at the highest dose of 900 mg QD for all analytes.

Peak (Cmax,ss) and overall (AUCtau) exposure to CMS121 and CMS121-C2 increased in a dose-proportional manner. For CMS121-C1, AUCtau was dose proportional while Cmax,ss was slightly greater than dose proportional, while for CMS121-C3, AUCtau was slightly less than dose proportional and Cmax,ss was dose proportional (Table 6a and 6b).

**Table 6a.** Summary of the Dose Proportionality Analysis of Plasma CMS121 Pharmacokinetic Parameters Following Multiple Oral Doses of 150 mg to 900 mg CMS121 QD (Part 2, MAD, Day 7)

| Pharmacokinetic Parameter | Estimate of Slope (b) | Standard Error | 95% Confidence Interval for Slope |
| --- | --- | --- | --- |
| AUC<sub>tau</sub> (ng h/mL) | 0.9949 | 0.1664 | 0.6499 - 1.3400 |
| C<sub>max,ss</sub> (ng/mL) | 0.9483 | 0.2001 | 0.5333 - 1.3633 |
| Parameters were ln-transformed prior to analysis.<br>The following statistical model using PROC MIXED of SAS was used to test dose proportionality:<br>Parameter = a * Dose<sup>b</sup> which is equivalent to ln(Parameter) = ln(a) + b[ln(Dose)]<br>Program: /CA32505/sas_prg/pksas/adam_intext_stats_dose.sas 28JUN2023 15:17 |  |  |  |

**Table 6b.** Summary of the Dose Proportionality Analysis of Plasma CMS121-C2 Pharmacokinetic Parameters Following Multiple Oral Doses of 150 mg to 900 mg CMS121 QD (Part 2, MAD, Day 7)

| Pharmacokinetic Parameter | Estimate of Slope (b) | Standard Error | 95% Confidence Interval for Slope |
| --- | --- | --- | --- |
| AUCtau (ng h/mL) | 0.8861 | 0.1609 | 0.5525 - 1.2197 |
| Cmax,ss (ng/mL) | 1.2959 | 0.1946 | 0.8922 - 1.6996 |
| Parameters were ln-transformed prior to analysis.<br>The following statistical model using PROC MIXED of SAS was used to test dose proportionality: $\text{Parameter} = a * \text{Dose}^b$ which is equivalent to $\ln(\text{Parameter}) = \ln(a) + b[\ln(\text{Dose})]$<br>Program: /CA32505/sas_prg/pksas/adam_intext_stats_dose.sas 28JUN2023 15:17 |  |  |  |

Urinary excretion of CMS121 metabolites was minimal, with the mean cumulative fraction excreted at 72 hours postdose on Day 7 being 0.411% or less for all doses.

#### Part 3, Age Effect

When compared to young subjects, elderly subjects had higher concentrations of CMS121 and its metabolites throughout the PK sampling interval on both Day 1 and Day 7 (Figure 4 and Table 7). Tmax tended to be similar for both groups, but CMS121 and its metabolites remained quantifiable for longer in elderly subjects. Likewise, the terminal elimination half-life of CMS121, CMS121-C2, and CMS121-C3 was considerably longer in elderly subjects.

**Table 7:** Summary of Plasma <u>CMS121</u> Pharmacokinetic Parameters Following Multiple Oral Doses of 600 mg CMS121 QD in Healthy Young or Elderly Adult Subjects (**Part 3, Elderly**)

| Day | Pharmacokinetic Parameters | Treatment M3 | Treatment E |
| --- | --- | --- | --- |
| 1 | AUC0-t (ng h/mL) | 27.14 (28.9) [n=6] | 63.90 (58.9) [n=6] |
|  | AUC0-24 (ng h/mL) | 27.18 (28.8) [n=6] | 64.00 (58.9) [n=6] |
|  | Cmax (ng/mL) | 9.220 (31.2) [n=6] | 18.85 (51.2) [n=6] |
|  | Tmax (h) | 2.005 (1.50, 4.00) [n=6] | 3.002 (1.51, 4.00) [n=6] |
|  | Tlast (h) | 23.333 (23.33, 23.36) [n=6] | 23.333 (23.33, 23.71) [n=6] |
| 7 | AUCtau (ng h/mL) | 32.21 (45.8) [n=6] | 95.90 (54.3) [n=6] |
|  | Cavg (ng/mL) | 1.342 (45.8) [n=6] | 3.996 (54.3) [n=6] |
|  | Cmax,ss (ng/mL) | 13.49 (62.7) [n=6] | 19.25 (59.4) [n=6] |
|  | Cmin,ss (ng/mL) | 0.1414 (59.4) [n=6] | 0.8498 (110.9) [n=6] |
|  | Tmax,ss (h) | 1.750 (1.00, 2.00) [n=6] | 1.999 (1.50, 3.13) [n=6] |
|  | Kel (1/h) | 0.06364 ± 0.013832 [n=3] | 0.06261 ± 0.057046 [n=5] |
|  | t½ (h) | 11.208 ± 2.1900 [n=3] | 22.249 ± 18.0119 [n=5] |
|  | CLss/F (L/h) | 20150 ± 8703.6 [n=6] | 6970 ± 3525.5 [n=6] |
|  | Vz,ss/F (L) | 285400 ± 86260 [n=3] | 243300 ± 263830 [n=5] |
|  | RA AUC | 1.222 ± 0.3387 [n=6] | 1.732 ± 1.2681 [n=6] |
|  | RA Cmax | 1.640 ± 1.0345 [n=6] | 1.159 ± 0.6878 [n=6] |
| Treatment M3: Multiple oral doses of 600 mg CMS121 once daily (QD) on Days 1 through 7<br>Treatment E: Multiple oral doses of 600 mg CMS121 once daily (QD) on Days 1 through 7 in healthy elderly adult subjects<br>AUCs, Cmax, Cmax,ss, Cmin,ss, and Cavg are presented as geometric mean and geometric CV%.<br>Tmax, Tmax,ss and Tlast are presented as median (minimum, maximum).<br>Other parameters are presented as arithmetic mean ± SD.<br>Program: /CA32505/sas_prg/pksas/adam_intext_pkparam.sas 15JUN2023 16:09 |  |  |  |

**Figure 4.**
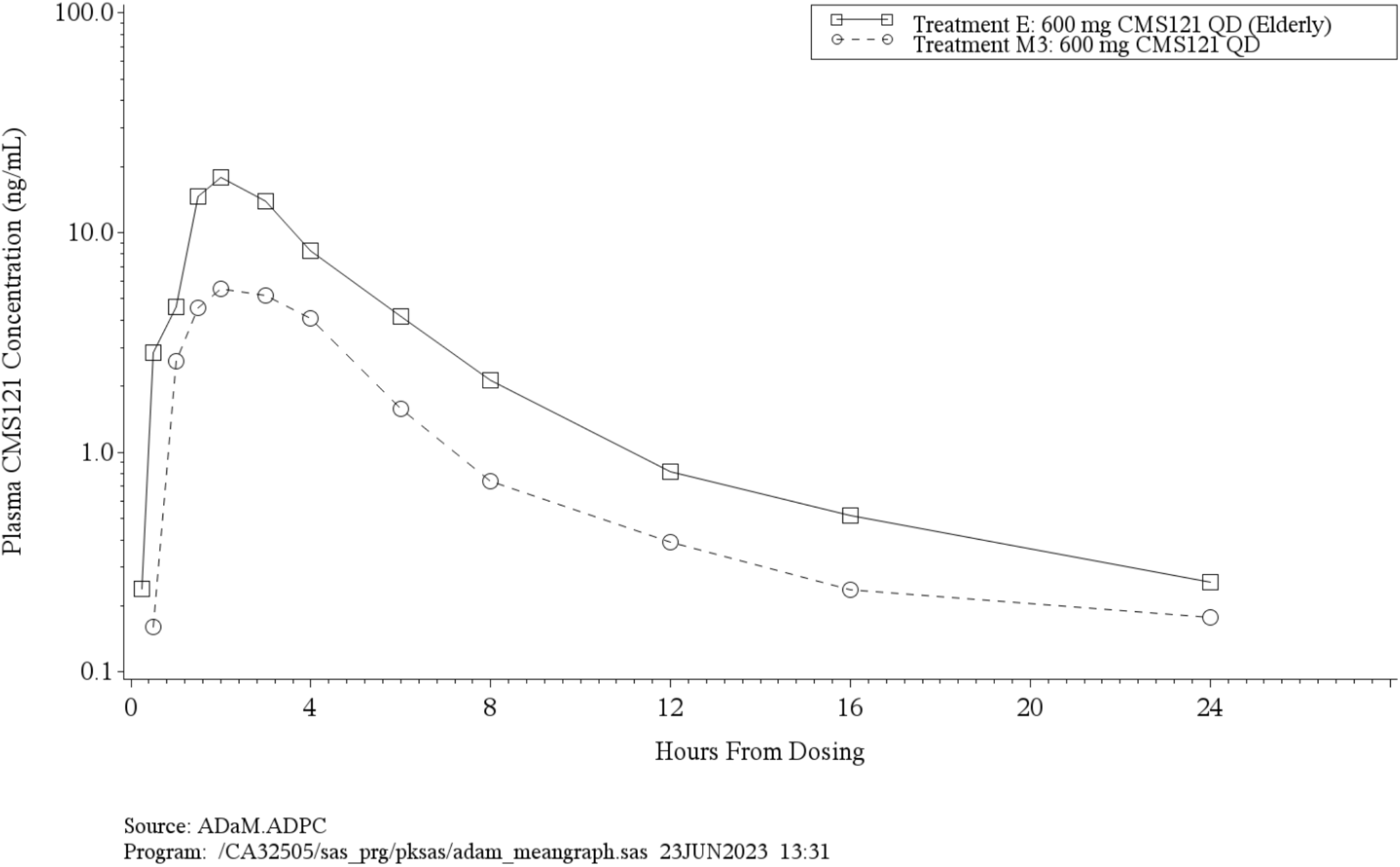
Arithmetic Mean <u>CMS121</u> Concentration Versus Time Profiles Following Multiple Oral Doses of 600 mg CMS121 QD in Healthy Young or Elderly Adult Subjects (Part 2, MAD, and Part 3, Age Effect, Day 1) (Semi-Log Scale)

Statistical comparisons indicated that peak (Cmax on Day 1) and overall (AUC0-t and AUC0-24 on Day 1, and AUCtau on Day 7) exposures to CMS121 and its metabolites were significantly higher in elderly subjects. However, this could not be concluded for Cmax,ss on Day 7 due to it being considerably more variable than the other parameters assessed (Table 8).

**Table 8:** Statistical Comparisons of Plasma <u>CMS121</u> Pharmacokinetic Parameters: 600 mg CMS121 QD (Elderly) Versus 600 mg CMS121 QD (**Part 2, MAD, and Part 3, Age Effect**)

|  |  | Treatment E (Test) |  | Treatment M3 (Reference) |  |  |  |  |
| --- | --- | --- | --- | --- | --- | --- | --- | --- |
| Parameter | Day | Geometric LSMs | n | Geometric LSMs | n | GMR (%) | 90% Confidence Interval | Inter-subject CV% |
| AUC0-t (ng h/mL) | 1 | 63.90 | 6 | 27.14 | 6 | 235.39 | 149.33 - 371.03 | 45.63 |
| AUC0-24 (ng h/mL) | 1 | 64.00 | 6 | 27.18 | 6 | 235.42 | 149.40 - 370.96 | 45.59 |
| Cmax (ng/mL) | 1 | 18.85 | 6 | 9.220 | 6 | 204.40 | 134.00 - 311.78 | 42.05 |
| AUCtau (ng h/mL) | 7 | 95.90 | 6 | 32.21 | 6 | 297.75 | 181.39 - 488.76 | 50.15 |
| Cmax,ss (ng/mL) | 7 | 19.25 | 6 | 13.49 | 6 | 142.70 | 79.18 - 257.18 | 61.05 |

**Treatment E:** Multiple oral doses of 600 mg CMS121 once daily (QD) on Days 1 through 7 in healthy elderly adult subjects (test)
**Treatment M3:** Multiple oral doses of 600 mg CMS121 once daily (QD) on Days 1 through 7 (reference)
Geometric least-squares means (LSMs) are calculated by exponentiating the LSMs derived from ANOVA.
Geometric Mean Ratio (GMR) = 100 x (test/reference)
Parameters were ln-transformed prior to analysis.
Inter-subject CV% was calculated as 100 x square root(exp[MSE]-1), where MSE = Residual variance from ANOVA.
Program: /CA32505/sas\_prg/pksas/adam\_intext\_statsmixed.sas 15JUN2023 16:07

While urinary excretion of CMS121 and its metabolite were still minimal in both groups, the cumulative fraction excreted at 72 hours postdose on Day 7 was markedly higher in elderly subjects, particularly for CMS121-C1 (0.191% in Elderly vs 0.060% in Young) and CMS121-C2 (1.054% vs. 0.275%). These results suggest that dose reductions may be required when CMS121 is administered to elderly subjects.

#### Part 4, Food Effect

When compared to the fasted state, food delayed the absorption of CMS121 and its metabolites as the peak concentration tended to occur at ∼4 hours in the fed state and 2-3 hours in the fasted state (Figure 5 and Table 9). Peak concentrations were also higher in the fed state for CMS121 and CMS121-C3. Statistical comparisons showed that overall (AUC0-t, but not AUC0-inf) and peak (Cmax) exposures to CMS121 and CMS121-C3 were higher in the fed state by approximately 50%, but there were no significant differences for CMS121-C1 and CMS121-C2 (Table 10).

**Table 9:**
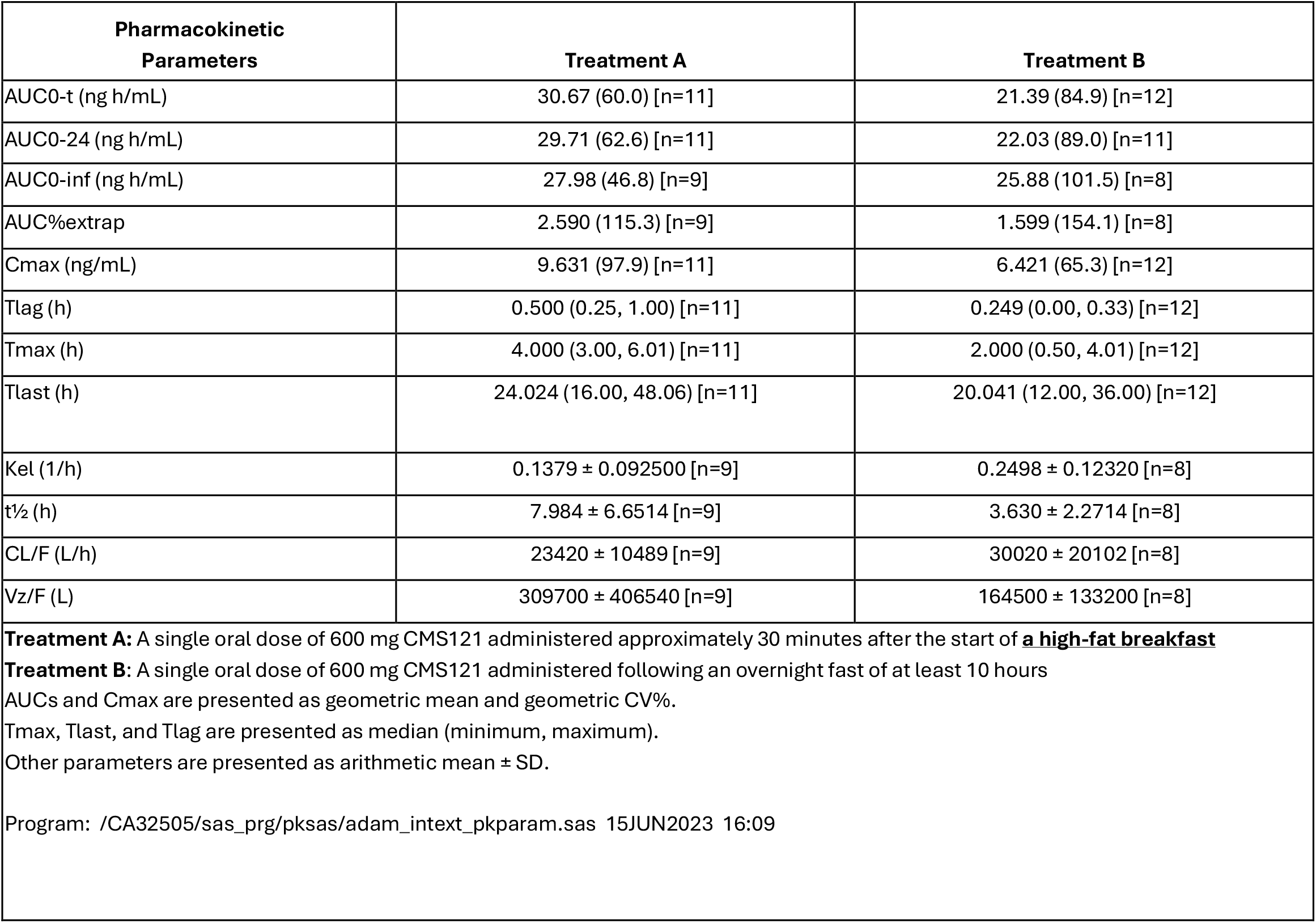
Summary of Plasma <u>CMS121</u> Pharmacokinetic Parameters Following a Single Oral Dose of 600 mg CMS121 Under Fed or Fasted Conditions (**Part 4, Food Effect**)

**Table 10:** Statistical Comparisons of Plasma <u>CMS121</u> Pharmacokinetic Parameters: 600 mg CMS121 (Fed) Vs 600 mg CMS121 (Fasted) (**Part 4, Food Effect**)

|  | Treatment A (Test) |  | Treatment B (Reference) |  |  |  |  |
| --- | --- | --- | --- | --- | --- | --- | --- |
| Parameter | Geometric LSMs | n | Geometric LSMs | n | GMR (%) | 90% Confidence Interval | Intra-subject CV% |
| AUC0-t (ng h/mL) | 31.65 | 11 | 21.39 | 12 | 147.92 | 111.30 - 196.58 | 37.78 |
| AUC0-inf (ng h/mL) | 29.10 | 9 | 22.62 | 8 | 128.64 | 84.51 - 195.82 | 39.83 |
| Cmax (ng/mL) | 9.662 | 11 | 6.421 | 12 | 150.48 | 111.73 - 202.65 | 39.71 |

**Treatment A:** A single oral dose of 600 mg CMS121 administered approximately 30 minutes after the start of a **high-fat breakfast (test)**
**Treatment B:** A single oral dose of 600 mg CMS121 administered following an overnight fast of at least 10 hours (**reference**)
Geometric least-squares means (LSMs) are calculated by exponentiating the LSMs derived from ANOVA.
Geometric Mean Ratio (GMR) = 100 x (test/reference)
Parameters were ln-transformed prior to analysis.
Intra-subject CV% was calculated as 100 x square root( $\exp[\text{MSE}] - 1$ ), where MSE = Residual variance from ANOVA.
Program: /CA32505/sas\_prg/pksas/adam\_intext\_statsmixed.sas 15JUN2023 16:07

**Figure 5.**
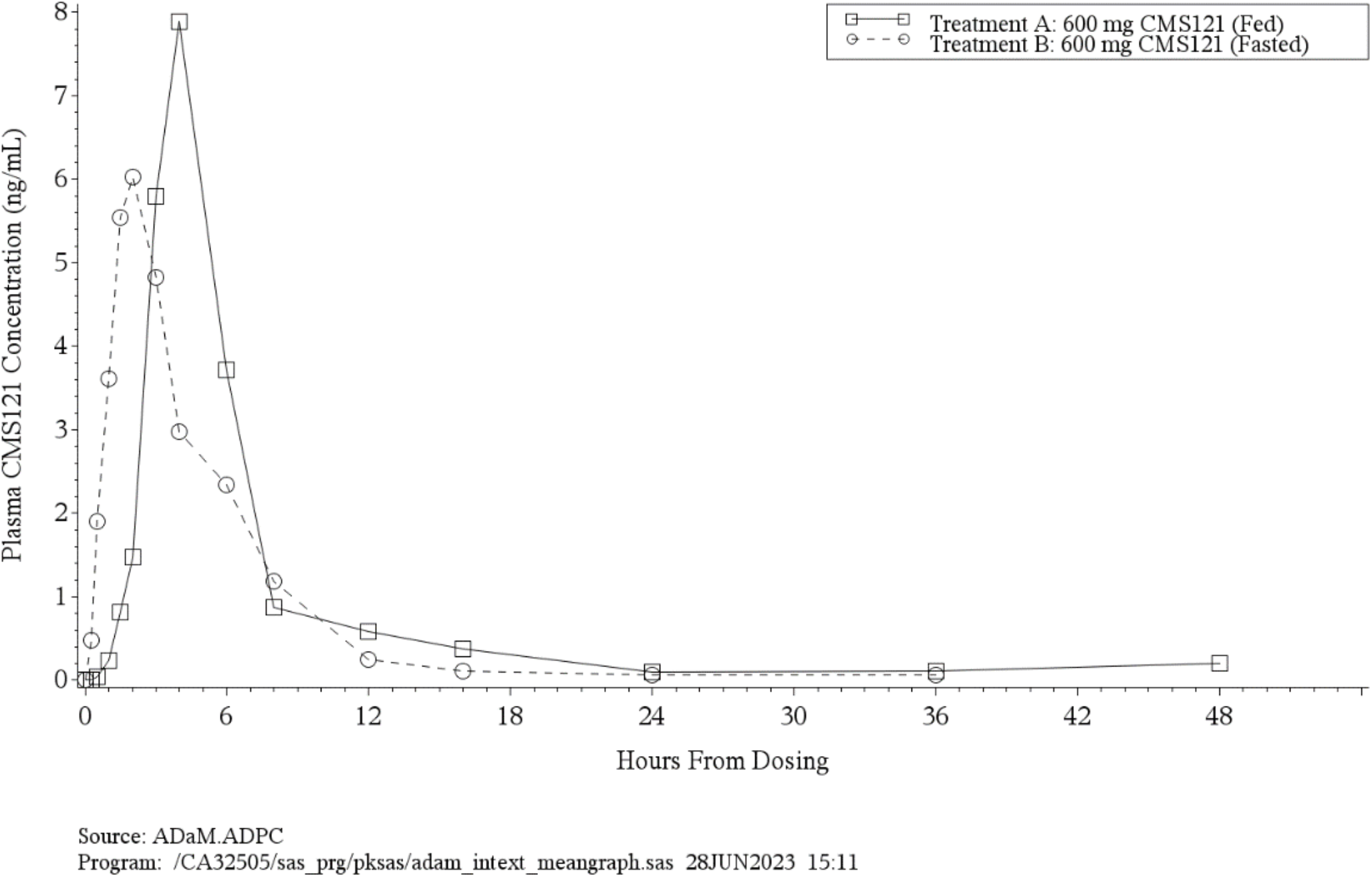
Arithmetic Mean <u>CMS121</u> Concentration Versus Time Profiles Following Single Oral Doses of 600 mg CMS121 Under Fed or Fasted Conditions (Part 4, Food Effect) (Linear Scale)

### Cardiodynamics

Overall, following single- and multiple-day dosing of CMS121, mean triplicate-average Holter monitor 12-lead ECG parameters remained within normal limits for all postdose time points and changes from baseline were generally small. Mean QTcF intervals were < 450 msec and changes from baseline were < 10 msec at all time points.

Following single-dose administration of up to 1800 mg of CMS121, statistical comparisons of QTcF change from baseline

## Discussion

The four phase 1 studies with CMS121 delineated the safety, tolerability, PK, and cardiodynamic profiles of this compound and food effect in healthy adult and elderly subjects.

Single ascending oral doses of CMS121 up to 1800 mg appeared to be safe and well tolerated by the healthy young adult subjects in this study. Multiple ascending oral doses of CMS121 up to 900 mg appeared to be safe and well tolerated by the healthy young adult subjects as well. Multiple oral doses of 600 mg CMS121 appeared to be safe and well tolerated by the healthy elderly subjects in this study. Single oral doses of 600 mg CMS121 administered to healthy young subjects under fed or fasting conditions appeared to be safe and well tolerated by the healthy young adult subjects in this study.

In summary, four studies constituting the Phase I program with the novel small molecule fisetin derivative, CMS121, assessed safety, PK, cardiodynamics, age-related and food effects in healthy adult subjects.

There were no deaths or SAEs during this study. One (1) subject in Part 3 (Elderly) missed a dose of study drug on Day 3 due to experiencing a dizziness AE and 1 subject in Part 4 (Food Effect) was discontinued due to a COVID-19 AE; both of these AEs were considered unrelated to the study drug by the PI. Across study parts, the majority of TEAEs were mild in severity and considered related to study product. There were no remarkable findings regarding clinical laboratory assessments, vital signs, physical examinations, or C-SSRS data in this study with respect to subject safety.

Oral administration of SD up to 1800 mg or RD up to 900 mg/day for 7 days was generally well tolerated, with concentration profiles for CMS121 and its metabolites peaking rapidly postdose, followed by a biphasic elimination profile. Peak (Cmax) and overall (AUCs) exposure to (dQTcF) between CMS121 and placebo were small, with negative time-matched differences at the majority of time points through Hour 24 (Table 11). Following multiple-day dosing, statistical comparisons of dQTcF between the 300 mg and 600 mg CMS121 dose levels and placebo exceeded 10 msec at several time points. Mean dQTcF differences for the 150 mg and 900 mg CMS121 QD dose levels compared to placebo were small at all time points (Table 12). The dQTcF that exceeded 10 msec at several time points might be due to the small sample size and large variations because the sample size was determined without statistical considerations.

**Table 11.** Error! No text of specified style in document. **Model Predicted ddQTcF and 90% CI at Mean Plasma CMS121 C**_**max**_ **by Treatment - Day 1 (Part 1, SAD) (C-QT Population)**

| **Treatment** | **Geometric Mean C<sub>max</sub>**<br>**(ng/mL)** | **Predicted ddQTcF**<br>**(msec)** | **90% Confidence Interval**<br>**(msec)** |
| --- | --- | --- | --- |
| S1 | 0.3437 | -1.847 | -4.239 - 0.545 |
| S2 | 1.231 | -1.852 | -4.241 - 0.538 |
| S3 | 3.070 | -1.861 | -4.247 - 0.525 |
| S4 | 5.923 | -1.876 | -4.264 - 0.511 |
| S5 | 29.01 | -1.998 | -4.673 - 0.677 |
| S6 | 20.30 | -1.952 | -4.464 - 0.561 |
| Treatment S1: A single oral dose of 50 mg CMS121<br>Treatment S2: A single oral dose of 150 mg CMS121<br>Treatment S3: A single oral dose of 300 mg CMS121<br>Treatment S4: A single oral dose of 600 mg CMS121<br>Treatment S5: A single oral dose of 1200 mg CMS121<br>Treatment S6: A single oral dose of 1800 mg CMS121<br>Program: /CA32505/sas_prg/pksas/cardio/intext_cqt_stats.sas 16JUN2023 10:19 |  |  |  |

**Table 12:** Model Predicted ddQTcF and 90% CI at Mean Plasma CMS121 C_max,ss_ by Treatment - Day 7 (Part 2, MAD) (C-QT Population)

| Treatment | Geometric Mean C<sub>max</sub><br>(ng/mL) | Predicted ddQTcF<br>(msec) | 90% Confidence Interval<br>(msec) |
| --- | --- | --- | --- |
| M1 | 2.193 | 4.857 | -3.063 - 12.776 |
| M2 | 5.627 | 5.173 | -2.763 - 13.109 |
| M3 | 13.49 | 5.898 | -2.181 - 13.977 |
| M4 | 10.22 | 5.596 | -2.405 - 13.598 |
| Treatment M1: 150 mg CMS121 QD<br>Treatment M2: 300 mg CMS121 QD<br>Treatment M3: 600 mg CMS121 QD<br>Treatment M4: 900 mg CMS121 QD<br>Program: /CA32505/sas_prg/pksas/cardio/sdtm_numlis.sas 22JUN2023 13:44 |  |  |  |

No subjects experienced postdose triplicate-average QTcF intervals exceeding 450 msec or QTcF interval changes from baseline ≥ 30 msec. There were no remarkable findings in the individual cardiodynamic assessments in this study.

CMS121 and CMS121-C1 metabolite increased in a slightly greater than dose-proportional manner. For CMS121-C2, dose proportionality was shown for AUC0-inf, but Cmax, AUC0-t, and AUC0-24 were slightly greater than dose proportional. CMS121 and its metabolites did not display any appreciable accumulation in total exposure over time. but accumulation was noted for peak concentrations, especially at the lower doses, for the parent, CMS121-C1, and CMS121-C2. Urinary excretion of CMS121 metabolites was minimal, with the mean molar-equivalent CMS121 cumulative fraction excreted at 72 hours postdose on Day 7 being 0.411% or less for all doses.

Multiple oral doses of 600 mg CMS121 appeared to be safe and well tolerated by the healthy elderly subjects. When compared to young subjects, elderly subjects had higher concentrations of CMS121 and its metabolites throughout the PK sampling interval on both Day 1 and Day 7, with all analytes remaining quantifiable for longer in elderly subjects. Likewise, the mean terminal elimination half-lives of CMS121, CMS121-C2, and CMS121-C3 were considerably longer in elderly subjects.

When compared to the fasted state, food delayed the absorption of CMS121 and the formation of its metabolites. Mean peak concentrations were also higher in the fed state for CMS121 and CMS121-C3.

Mean QTcF intervals were < 450 msec and changes from baseline were < 10 msec at all time points across CMS121 dose levels and placebo following single-day dosing up to 1800 mg and multiple-day dosing of 150 mg. Mean Holter monitor 12-lead ECG parameters (QTcF, HR, RR, PR, and QRS) remained within normal limits for all postdose time points and changes from baseline were generally small. There were no individual categorical outliers with an absolute QTcF > 450 msec or QTcF interval change from baseline ≥ 30 msec. There were no significant findings on the cardiodynamic parameters following single doses of CMS121 up to 1800 mg and multiple doses up to 900 mg QD.

These dose ranges will be utilised to further explore the safety, PK and potential for therapeutic effect of CMS121 in the management of AD in planned Phase 2 clinical studies.

## Data Availability

All data produced in the present study are available upon reasonable request to the authors

## Acknowledgements

The authors wish to thank Celerion, Inc. for their contribution to the clinical studies. Additionally, the authors are grateful to Erin Castelloe, MD for her contributions to the study design and monitoring of the clinical studies.

## Supporting Information

Additional supporting information may be found in the online version of this article at the publisher’s web-site.

## Safety Tables and Figures

## Pharmacokinetic Tables and Figures

**Summary of Plasma Pharmacokinetic Parameters for CMS121 and the Primary Metabolite (CMS121-C2) Across Study Groups**

**Figures: Plasma Concentration vs timecourse profiles**

## Cardiodynamic Safety Tables

